# Real-time analysis of a mass vaccination effort confirms the safety of FDA-authorized mRNA vaccines for COVID-19 from Moderna and Pfizer/BioNtech

**DOI:** 10.1101/2021.02.20.21252134

**Authors:** Reid McMurry, Patrick Lenehan, Samir Awasthi, Eli Silvert, Arjun Puranik, Colin Pawlowski, AJ Venkatakrishnan, Praveen Anand, Vineet Agarwal, John C. O’Horo, Gregory J. Gores, Amy W. Williams, Andrew D. Badley, John Halamka, Abinash Virk, Melanie D. Swift, Katie Carlson, Deeksha Doddahonnaiah, Anna Metzger, Nikhil Kayal, Gabi Berner, Eshwan Ramudu, Corinne Carpenter, Tyler Wagner, Ajit Rajasekharan, Venky Soundararajan

## Abstract

As the COVID-19 vaccination campaign unfolds as one of the most rapid and widespread in history, it is important to continuously assess the real-world safety of the FDA-authorized vaccines. Curation from large-scale electronic health records (EHRs) allows for near real-time safety evaluations that were not previously possible. Here, we advance context- and sentiment-aware deep neural networks over the multi-state Mayo Clinic enterprise (Minnesota, Arizona, Florida, Wisconsin) for automatically curating the adverse effects mentioned by healthcare providers in over 108,000 EHR clinical notes between December 1^st^ 2020 and February 8^th^ 2021. We retrospectively compared the clinical notes of 31,029 individuals who received at least one dose of the Pfizer/BioNTech or Moderna mRNA vaccine to those of 30,933 unvaccinated individuals who were propensity matched by demographics, residential location, and history of prior SARS-CoV-2 testing. We find that vaccinated and unvaccinated individuals were seen in the clinic at similar rates within 21 days of the first or second actual or assigned vaccination date (first dose Odds Ratio = 1.14, 95% CI: 1.10-1.18; second dose Odds Ratio = 0.91, 95% CI: 0.86-0.96). Further, the incidence rates of all surveyed adverse effects were similar or lower in vaccinated individuals compared to unvaccinated individuals after either vaccine dose, although myalgia was modestly increased within 7 days of the second dose when considering only pairs of matched individuals who each had at least one clinical note in this time window (Incidence Rate Ratio = 2.5, 95% CI: 1.1-6.7). Finally, the most frequently documented adverse effects within 7 days of each vaccine dose were fatigue (Dose 1: 1.75%, Dose 2: 1.18%), nausea (Dose 1: 1.03%, Dose 2: 0.84%), myalgia (Dose 1: 0.41%; Dose 2: 0.43%), diarrhea (Dose 1: 0.65%; Dose 2: 0.45%), arthralgia (Dose 1: 0.64%; Dose 2: 0.57%), erythema (Dose 1: 0.56%; Dose 2: 0.44%), vomiting (Dose 1: 0.44%, Dose 2: 0.29%) and fever (Dose 1: 0.21%; Dose 2: 0.18%). These frequencies of adverse event documentation in EHR notes are 2.1 times (95% CI: [1.5, 3.0]) to 1500 times (95% CI: [670, 2800]) lower than the frequencies of adverse events recorded via active solicitation during clinical trials or post-marketing surveillance, with headache after second vaccination showing the highest ratio of trial reporting to EHR documentation. This rapid and timely analysis of EHR notes from 31,029 vaccinated individuals highlights the rarity of vaccine-associated adverse effects requiring clinical attention and reaffirms the tolerability of the FDA-authorized COVID-19 vaccines in practice.

## Introduction

Following their Emergency Use Authorizations by the Food and Drug Administration (FDA) in December of 2020, more than 52 million doses of BNT162b2 (Pfizer/BioNTech) and mRNA-1273 (Moderna) COVID-19 vaccines have been administered in the United States^1–3^. Phase 3 trials demonstrate strong efficacy and favorable safety profiles for these vaccines in the cohorts studied. Specifically, the trials showed 95.0% (95% CI, 90.3 to 97.6) and 94.1% (95% CI, 89.3 to 96.8%) efficacy for BNT162b2 and mRNA-1273 respectively. While self-resolving mild to moderate adverse effects were common in vaccinated participants, serious adverse effects occurred rarely and with a frequency comparable to placebo^4,5^. Local adverse effects of any severity reported in these trials included injection site pain (84.1-92%), injection site swelling (10.5-70%), and injection site erythema (9.5-14.6%). Systemic effects of any severity included fatigue (62.9-70%), headache (55.1-64.7%), myalgia (38.3-61.5%), arthralgia (23.6-46.4%), chills (31.9-45.4%), fever (14.2-15.5%), and nausea/vomiting (1.1-23.0%)^6,7^.

Consistent with CDC recommendations, early vaccination efforts (Phase 1a) in the United States have targeted healthcare workers, residents and staff at long term care facilities, who are at elevated risk for COVID-19 relative to the general public^8–11^. As the vaccines continue to be administered more broadly, it will be critical to continuously evaluate real world safety and efficacy data from all those who have received these vaccines. This approach may validate existing findings or highlight differences in the larger population compared to the clinical trial experience with respect to vaccine efficacy and adverse effects. Of note, the populations undergoing vaccination may differ meaningfully from the trial populations. For example, healthcare workers and long term care residents receiving the first wave of vaccines in Phase 1a likely comprised a small fraction of the Phase 3 trial populations. Additionally, some pregnant women are receiving COVID-19 vaccines in the real world^12^, while they were excluded from both trials^4,5^. Finally, continuous monitoring will help to identify and better quantify the frequency of rare severe adverse effects such as anaphylaxis, which was widely reported but only observed in a small number of individuals after approval of both vaccines^13–17^.

As part of the Vaccine Adverse Events Reporting System (VAERS) maintained by the FDA, V-safe has recently been established to specifically monitor the safety profiles of COVID-19 vaccines as they are administered to the public^18^. V-safe is a voluntary program into which vaccinated individuals can enroll. Once enrolled, participants receive frequent reminders to complete surveys which document their side effects electronically. Thus, this program will create an excellent resource of clinical trial-like safety data derived from significantly larger and more diverse patient populations than those who were enrolled in the Phase 1/2 or Phase 3 trials.

An important complementary approach to post-authorization surveillance of vaccine efficacy and safety is via the real-time analysis of patient data stored in electronic health record (EHR) systems. We have previously developed and described augmented curation methods to rapidly create and compare cohorts of COVID-19 patients within a large EHR system, and have recently applied these methods to assess the real world efficacy of both BNT162b2 and mRNA-1273 in over 31,000 individuals receiving these vaccines at the Mayo Clinic and associated health system^19–23^. Here we expand on these efforts to study the adverse effects experienced by individuals and documented in the EHR after COVID-19 vaccination in the clinical environment.

It should be noted that the monitoring of vaccine-associated adverse effects in clinical trials or post-marketing surveillance efforts like V-safe is quite different from such monitoring in routine clinical practice. In clinical trials, participants are aware that they are receiving an experimental product, and adverse effect reporting is encouraged or solicited. Similarly, as was previously noted, V-safe participants complete frequent surveys to document their side effects after vaccination. On the other hand, individuals receiving a COVID-19 vaccine during the mass vaccination campaign are informed of adverse effects that are likely to be experienced and can even be discouraged from seeking medical attention unless the symptom is particularly severe. Thus, adverse effects are likely to be captured only in the EHR of individuals who experience symptoms which are severe or persistent enough to warrant a return to clinic, or who happen to have a previously scheduled routine clinical visit in the post vaccination time period. Accordingly, the intention of our analysis is not to determine whether real world data recapitulates the adverse effect frequencies reported in prior trials. Instead, it is to (1) establish the rates at which individuals actually report potential vaccine-associated adverse effects to health care practitioners (HCPs) in several defined time intervals after vaccination, and (2) determine whether these rates of adverse effect reporting are unexpectedly high.

To address the latter point, it is critical to establish the baseline frequency at which each adverse effect is expected to be documented in the clinical notes of our vaccinated cohort. We thus built upon our previous vaccine efficacy analysis which employed a 1-to-1 propensity matching procedure to derive a cohort of unvaccinated individuals who are balanced for demographic factors, residential location, and history of SARS-CoV-2 PCR testing (see **Methods**)^24^. Here we curated the clinical notes of these vaccinated and matched unvaccinated individuals over several time periods to define the frequencies of various symptoms of interest. In our analyses, the propensity matched group serves a purpose similar to that of the placebo arm in clinical trial safety assessments, allowing us to contextualize and better interpret the absolute rates of adverse effects documented in the notes of vaccinated individuals.

We find that individuals undergoing COVID-19 vaccination do not return to the clinic, including the emergency department, at unexpectedly high rates in the 7, 14, or 21 days following either vaccine dose. Further, the rates of adverse effects (headache, myalgia, arthralgia, fatigue, fever, diarrhea, nausea/vomiting, anaphylaxis, facial paralysis, lymphadenopathy, and erythema) documented in EHR notes over these time intervals are generally not higher in vaccinated individuals compared to the unvaccinated control cohort. The absolute rates of adverse effect documentation in the EHR are markedly lower than the rates observed in settings of solicited reporting (i.e. clinical trials, post-marketing surveillance). This study supports the safety and tolerability of BNT162b2 and mRNA-1273 in practice, further strengthening the case for the rapid and broad distribution of vaccines to the public.

## Methods

### Study design, setting and population

This is a retrospective study of individuals who underwent polymerase chain reaction (PCR) testing for suspected SARS-CoV-2 infection at the Mayo Clinic and hospitals affiliated with the Mayo Clinic Health System. This study was reviewed by the Mayo Clinic Institutional Review Board (IRB) and determined to be exempt from the requirement for IRB approval (45 CFR 46.104d, category 4). Subjects were excluded if they did not have a research authorization on file.

The cohorts of vaccinated and unvaccinated individuals eligible for inclusion in this study are identical to the cohorts considered in a previous analysis: “FDA-authorized COVID-19 vaccines are effective per real-world evidence synthesized across a multi-state health system”^24^. In total, there were 507,525 individuals in the Mayo electronic health record (EHR) database who received a PCR test between February 15, 2020 and February 8, 2021. To obtain the eligible study population, we defined the following inclusion criteria: (1) at least 18 years old; (2) no positive SARS-CoV-2 PCR test before December 1, 2020; (3) resides in a locale (based on Zip code) with at least 25 individuals who have received BNT162b2 or mRNA-1273. This population included 249,708 individuals, of whom 31,623 have received BNT162b2 or mRNA-1273 and 218,085 have no record of COVID-19 vaccination. Vaccinated individuals who had tested positive for SARS-CoV-2 by PCR between December 1, 2020 and the date of their first vaccine dose were excluded, resulting in 31,299 individuals. Individuals with zero follow-up days after vaccination (i.e. those who received the first vaccine dose on the date of data collection) were also excluded, leaving 31,069 individuals in the final vaccinated cohort.

The propensity matched unvaccinated cohort was selected from the previously derived set of 218,085 unvaccinated individuals. The purpose of this cohort was to establish the baseline frequency of EHR documentation for each adverse effect of interest in a cohort which is clinically similar to our vaccinated cohort. These baseline, or expected, frequencies can then be compared to the observed frequencies to determine whether or not these adverse effects are reported at unexpectedly high rates among patients receiving a COVID-19 vaccine.

From these cohorts of 31,069 vaccinated and propensity matched unvaccinated individuals, we removed individuals who received a positive SARS-CoV-2 PCR test within the first four days after their first actual or assigned vaccine dose (see below). This filtering step was done because it is known that the onset of disease associated symptoms, some of which overlap with vaccine side effects, can precede the confirmed diagnosis of COVID-19 by PCR^25^. Accordingly, the cause of symptoms observed during this time period for such patients would be ambiguous, as they could reasonably stem from vaccination or infection. After this filter, there were 31,029 and 30,933 individuals in the final vaccinated and unvaccinated cohorts, respectively.

The selection algorithm and its associated counts are summarized in **Figure 1**. More details on the propensity score matching procedure are provided in the prior manuscript^24^. This includes a table showing that the 1-to-1 matched cohorts (n = 31,069 each) were indeed balanced for all covariates included in propensity score matching (standardized mean difference < 0.1 for each one) and figures illustrating the distributions of age and total follow-up time between the first vaccine dose and the end of the data collection period.

**Figure 1.**
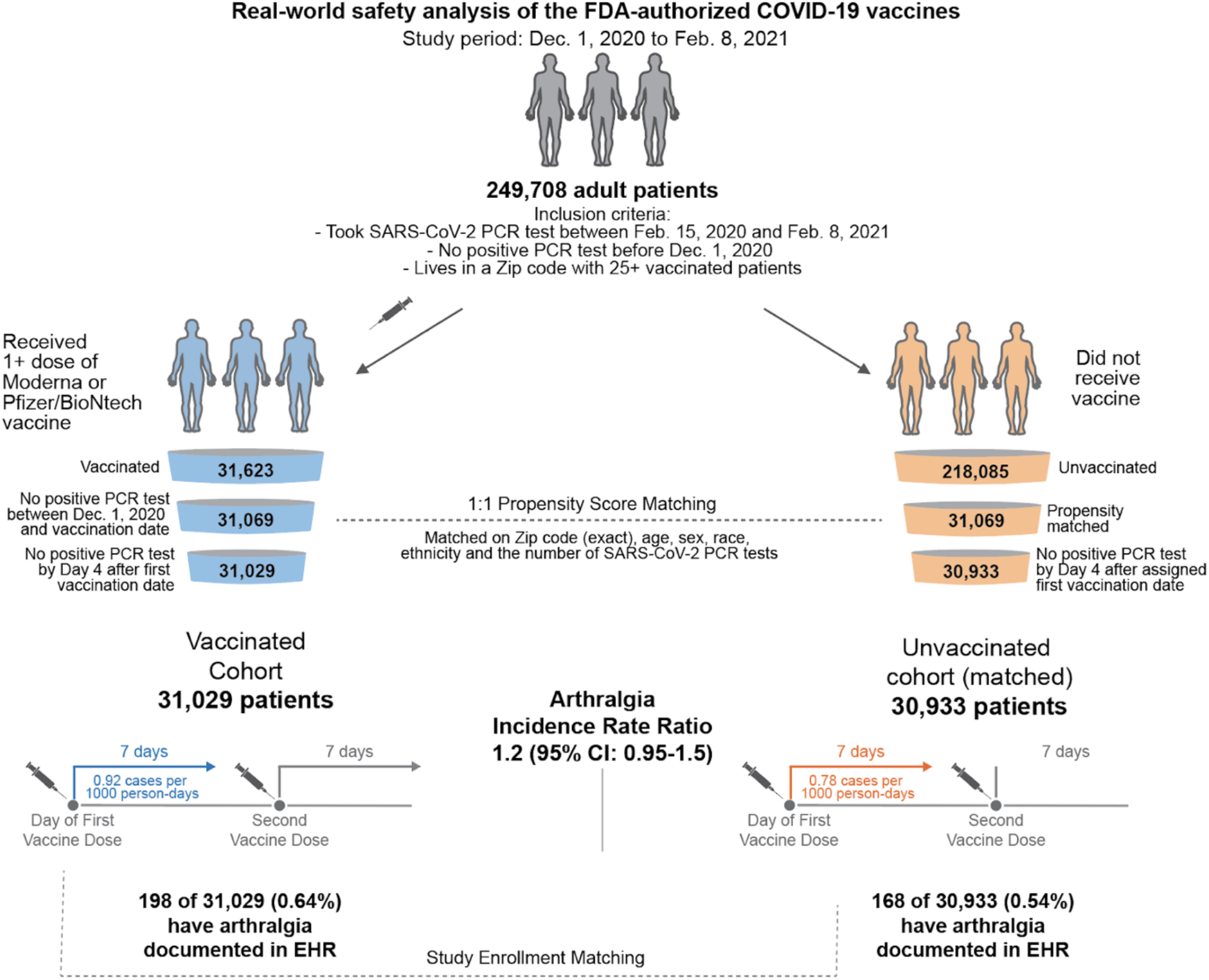
Schematic illustration of participant selection and study design. A vaccinated cohort and 1-to-1 propensity matched unvaccinated cohort (n = 31,069 each) were previously derived to assess vaccination efficacy in a separate study. With the additional specification that an individual must not have had a positive SARS-CoV-2 PCR test with the first four days after their actual or assigned vaccination date, we were left with 31,029 vaccinated individuals and 30,933 unvaccinated individuals for this safety analysis. For each cohort, the incidence rates of several adverse effects (e.g. arthralgia) were calculated for the seven days following the first dose and, separately, for the seven days following the second dose. For a given adverse effect, the incidence rate ratio and its corresponding 95% confidence interval (CI) were calculated to determine whether one cohort was more likely to experience the event than the other. Incidence rates and incidence rate ratios were also calculated for the 14 and 21 days following each vaccine dose.

### Definition of time intervals for safety analyses

For each vaccinated individual, we defined the date of their first vaccine dose as Day V_1_ and the date of their second vaccine dose as Day V_2_. In the Results section, these are referred to as “actual” dates of vaccination. For each unvaccinated individual, Day V_1_ and Day V_2_ were designated as identical to their matched vaccinated individual. In the Results section, these are referred to as “assigned” dates of vaccination. Individuals were considered for the post-second dose analyses if they (1) were part of the initial cohorts, (2) received a second vaccine dose (vaccinated cohort) or were matched to an individual who received a second vaccine dose (unvaccinated cohort), and (3) were not diagnosed with COVID-19 before Day V_2_ or within the first four days after Day V_2_. The third criterion was applied for a similar reason as explained above for the initial cohorts: if an individual tested positive for SARS-CoV-2 on or before the fourth day after the second vaccination, then any symptoms observed after the second actual or assigned vaccination date would have an ambiguous cause.

### Definition of adverse effects of interest

The adverse effects considered were primarily derived from those assessed in Phase III trials of BNT162b2 and mRNA-1273^4,5^, including fatigue, fever, chills, myalgia, arthralgia, headache, lymphadenopathy, erythema, diarrhea, vomiting, and local pain and swelling. We also included anaphylaxis and facial paralysis (Bell’s palsy), as these rare events have been reported in individuals receiving COVID-19 vaccines as well. Each adverse effect was mapped to a set of synonyms intended to capture the most common ways that a given phenotype would be referenced in the context of a clinical note.

### Curation of adverse effects from clinical notes

To curate the adverse effects experienced by each patient from the electronic health record, we used a BERT-based neural network model^26^ to classify the sentiment for the phenotypes (described above) mentioned in the clinical notes. Specifically, this classification model categorizes phenotype-containing sentences into one of four categories: (1) confirmed diagnosis, (2) ruled-out diagnosis, (3) possibility of disease, and (4) alternate context (e.g. family history). This classification model was trained on 18,500 sentences and has shown an out-of-sample accuracy of 93.6% with precision and recall scores above 95%^25^. For each individual, we applied the sentiment model to the clinical notes in the Mayo Clinic electronic health record during our defined intervals of interest for each individual: (1) Day V_1_ to 7, 14, or 21 days after Day V_1_, and (3) Day V_2_ to 7, 14, or 21 days after Day V_2_. For each phenotype, we identified the first date on which the given individual had at least one sentence in which the phenotype was categorized as “confirmed diagnosis” with a confidence score of at least 90%. For the severe phenotype anaphylaxis, each such sentence was manually reviewed to verify the positive sentiment (i.e. confirmed diagnosis) and to assess the tense of this sentiment (i.e. past vs. present). Only sentences which confirm a present diagnosis were used to count anaphylaxis events in this study.

### Evaluating rates of return to clinic after vaccination

To evaluate the likelihood of returning to the clinic after vaccination, we counted the number of individuals who had at least one clinical note in the 7, 14, and 21 days after Day V_1_ and Day V_2_. The fraction of individuals with clinical follow-up was calculated as the number of individuals with at least clinical note in the time window divided by the total number of individuals in each group. The difference in clinical follow-up rates was assessed by calculating the odds ratio (OR) along with its corresponding 95% confidence interval (CI). The null hypothesis was that the OR falls between 0.91 and 1.1, inclusive (i.e. the larger rate is at most 10% larger than the smaller rate); thus, an OR was considered significant if the upper bound of the 95% CI was less than 0.91 or the lower bound of the 95% CI was greater than 1.1.

To evaluate emergent clinical follow-up, we compared the number and percentage of vaccinated and unvaccinated individuals who contributed at least one Emergency Department (ED) note in the 1, 7, 14, or 21 days after each actual or assigned vaccine administration date. The odds ratio and corresponding 95% CI was calculated for each time window, with the null hypothesis stating that the OR falls between 0.91 and 1.1, inclusive (as above). We also determined the total number of ED notes from vaccinated and unvaccinated individuals in these same time windows, along with the fraction of ED notes relative to all clinical notes. The difference in the fraction of ED notes was assessed by computing an odds ratio and corresponding 95% CI, with the null hypothesis stating that the OR falls between 0.91 and 1.1, inclusive. ED notes included those under the following categories in the EHR system: ED Notes, ED Procedure Note, ED Provider Notes, ED Provider Triage Note, ED Re-evaluation Note, and ED Triage Notes. All odds ratios and 95% confidence intervals were calculated using the fisher.test function in R version 4.0.3^27^.

### Evaluating vaccine adverse effects among the total cohorts

To evaluate adverse effects associated with receiving a COVID-19 vaccine in the clinical setting, we compared the two populations described above and summarized in **Figure 1**: (1) 31,029 individuals with follow-up who received BNT162b2 or mRNA-1273 and did not have a positive SARS-CoV-2 PCR test prior to vaccination (“vaccinated”), and (2) 30,933 propensity matched individuals who have never received a COVID-19 vaccine and did not have a positive SARS-CoV-2 PCR test before the first vaccination date of their matched individual (“unvaccinated”).

The association of a given adverse effect to vaccination status was assessed by computing the incidence rate ratio (IRR) of the vaccinated and unvaccinated cohorts during several defined time intervals. Specifically, we evaluated adverse effects which were documented in clinical notes within 7, 14, or 21 days of receiving the first vaccine dose (Day V_1_ to 7, 14, or 21 days after Day V_1_) or the second vaccine dose (Day V_2_ to 7, 14, or 21 days after Day V_2_). For each cohort in a defined time period, incidence rates were calculated as the number of individuals experiencing the given adverse effect in that time period divided by the total number of at risk person-days contributed in that time period. For each individual, at-risk person-days are defined as the number of days from the start of the time period until the day on which the individual experienced the adverse effect or died, or four days prior to testing positive for SARS-CoV-2. The IRR was calculated as the incidence rate of the vaccinated cohort divided by the incidence rate of the unvaccinated cohort, and its 95% CI was computed using an exact approach described previously^28^. The IRR was considered to be statistically significant if the 95% CI did not include 1.

### Comparison of adverse effect rates per EHR versus clinical trial and V-safe reporting

We aimed to compare the percentage of patients with an EHR-documented occurrence of each adverse event to the percentage of patients who reported the corresponding adverse event in the Phase 3 trials of BNT162b2 and mRNA-1273, and in V-safe (a voluntary and active post-marketing surveillance effort to assess the safety of COVID-19 vaccines)^4,5^. To do so, we extracted the number of patients reporting each adverse effect (of any severity level) within 7 days of the first or second vaccine doses from the clinical trial Solicited Safety Set (mRNA-1273; N_Dose 1_ = 14,808, N_Dose 2_ = 14,733) or Reactogenicity Subset (BNT162b2; N_Dose 1_ = 4,093, N_Dose 2_ = 3,758). We also extracted the percentage of patients reporting each adverse effect through January 14, 2021 in response to daily text message-based health screenings as part of V-safe for the week following each dose of vaccination^29^. We compared these clinical trial and post-marketing surveillance adverse effect reporting rates to the rates of EHR-documented adverse effects within 7 days of the first or second doses. Reporting-to-Documentation ratios were calculated for each adverse effect in each time interval by dividing the 7 day reporting rate in the given clinical trial (or V-safe) by the 7 day EHR documentation rate in our cohort. 95% confidence intervals around these ratios were computed with a standard delta-method-based approach^30^.

### Evaluating vaccine adverse effects in individuals who returned to clinic

Because only a fraction of individuals in the vaccinated and unvaccinated cohorts contributed clinical notes during the defined intervals after their actual or assigned vaccination dates, we also assessed the rates of adverse effects documented in the EHR specifically among the individuals who had returned to clinic. To select the cohorts for this analysis, we considered any pair of matched vaccinated and unvaccinated individuals who both contributed at least one clinical note which contained at least one phenotype term in the 7, 14, or 21 days after the actual or assigned date of first or second vaccination. The number of such matched pairs in each time interval is provided in **Table S1**. For each time interval, we then computed incidence rates for each adverse effect in the vaccinated and unvaccinated cohorts, along with the incidence rate ratios and 95% CI as described above. The IRR was considered to be statistically significant if the 95% CI did not include 1.

## Results

### Vaccinated individuals do not return to the clinic or emergency department at unexpectedly high rates after either vaccine dose

To assess rates of clinical follow-up after study enrollment, we compared the number of vaccinated and unvaccinated individuals with EHR notes within 7, 14, or 21 days of each actual or assigned vaccination date. In the 7 days after the first vaccine dose, 6,081 of the 31,029 (19.6%) vaccinated individuals had at least one EHR note, compared to 5,653 of 30,933 (18.3%) unvaccinated individuals (Odds Ratio = 1.09; 95% CI: [1.05-1.13]) (**Table 1**). This difference was not considered significant as the lower bound of the 95% CI was less than 1.1 (see **Methods**). The rates of return to clinic were also similar between these cohorts within 14 and 21 days of the first dose (Odds Ratio_14 Days_ = 1.13 [1.09-1.17]; Odds Ratio_21 Days_ = 1.14 [1.10-1.18]) (**Table 1**).

**Table 1.** Rates of return to clinic in vaccinated and unvaccinated individuals after first vaccination. The first two columns give the numbers and percent of individuals in each group who had at least one phenotype-containing EHR note with 7, 14, or 21 days of the actual or assigned first vaccination date. To assess the magnitude and significance of difference between the follow-up rates, the odds ratio (OR) and corresponding 95% CI are shown. With the null hypothesis that the OR falls between 0.91 and 1.1, a difference was considered significant if the upper bound of 95% CI was less than 0.91 or the lower bound of the 95% CI was greater than 1.1.

|  | **Vaccinated**<br>**individuals with at**<br>**least one note (%)**<br>**[n = 31,029]** | **Unvaccinated**<br>**individuals with at**<br>**least one note (%)**<br>**[n = 30,933]** | **Odds Ratio**<br>**(95% CI)** |
| --- | --- | --- | --- |
| **Within 7 days of**<br>**first actual or**<br>**assigned**<br>**vaccination date** | 6,081<br>(19.6%) | 5,653<br>(18.3%) | 1.09<br>(1.05-1.13) |
| **Within 14 days of**<br>**first actual or**<br>**assigned**<br>**vaccination date** | 8,743<br>(28.2%) | 7,990<br>(25.8%) | 1.13<br>(1.09-1.17) |
| **Within 21 days of**<br>**first actual or**<br>**assigned**<br>**vaccination date** | 10,438<br>(33.6%) | 9,537<br>(30.8%) | 1.14<br>(1.10-1.18) |

Within 7 days of the second dose, 2,496 of 16,982 (14.7%) vaccinated individuals contributed EHR notes, compared to 2,630 (15.8%) of 16,623 unvaccinated individuals (Odds Ratio = 0.92 [0.86-0.97) (**Table 2**). This difference was not considered significant as the upper bound of the 95% CI was greater than 0.91 (see **Methods**). Similarly, the rates of return to clinic were comparable between these cohorts within 14 and 21 days of the second dose (Odds Ratio_14 Days_ = 0.90 [0.85-0.95]; Odds Ratio_21 Days_ = 0.91 [0.86-0.96]) (**Table 2**).

**Table 2.**
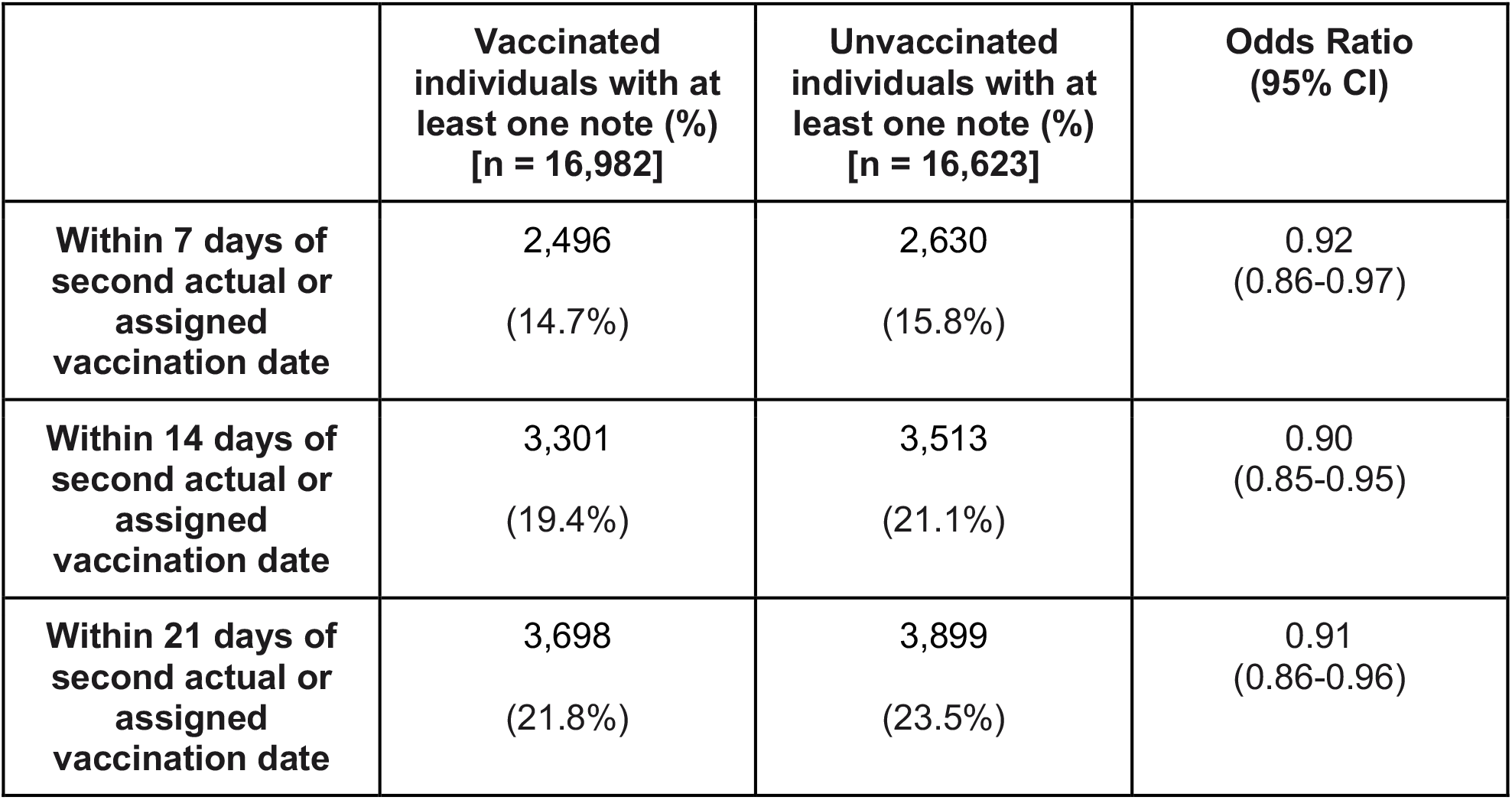
Rates of return to clinic in vaccinated and unvaccinated individuals after second vaccination. The first two columns give the numbers and percent of individuals in each group who had at least one phenotype-containing EHR note with 7, 14, or 21 days of the actual or assigned first vaccination date. To assess the difference in follow-up rates between groups, the odds ratio (OR) and corresponding 95% CI are shown. With the null hypothesis that the OR falls between 0.91 and 1.1, a difference was considered significant if the upper bound of 95% CI was less than 0.91 or the lower bound of the 95% CI was greater than 1.1.

|  | **Vaccinated**<br>**individuals with at**<br>**least one note (%)**<br>**[n = 16,982]** | **Unvaccinated**<br>**individuals with at**<br>**least one note (%)**<br>**[n = 16,623]** | **Odds Ratio**<br>**(95% CI)** |
| --- | --- | --- | --- |
| **Within 7 days of**<br>**second actual or**<br>**assigned**<br>**vaccination date** | 2,496<br>(14.7%) | 2,630<br>(15.8%) | 0.92<br>(0.86-0.97) |
| **Within 14 days of**<br>**second actual or**<br>**assigned**<br>**vaccination date** | 3,301<br>(19.4%) | 3,513<br>(21.1%) | 0.90<br>(0.85-0.95) |
| **Within 21 days of**<br>**second actual or**<br>**assigned**<br>**vaccination date** | 3,698<br>(21.8%) | 3,899<br>(23.5%) | 0.91<br>(0.86-0.96) |

To specifically assess the rates of emergent clinical follow-up, we also compared the number of emergency department (ED) EHR notes contributed by each group. The vaccinated cohort contributed a similar or lower number of ED notes than the unvaccinated cohort in the 1, 7, 14, and 21 days after both the first and second actual or assigned vaccination dates (**Tables S2-S3**). Consistent with this, fewer vaccinated than unvaccinated individuals contributed at least one ED EHR note within any of these time intervals (**Tables S4-S5**).

In summary, these findings show that individuals receiving COVID-19 vaccines do not tend to return to the clinic, including the emergency department, in the subsequent weeks at higher-than-expected rates, which suggests favorable tolerability of these vaccines.

### Documented vaccine associated adverse effects in EHR notes are rare compared to the frequencies reported in clinical trials

Among the 31,029 and 30,933 individuals in the vaccinated and unvaccinated cohorts, the most commonly documented symptoms in the 7 days after the first vaccine dose included fatigue (542 [1.75%] vaccinated individuals; 592 [1.91%] unvaccinated individuals), nausea (321 [1.03%]; 493 [1.59%]), diarrhea (203 [0.65%]; 247 [0.80%]), myalgia (128 [0.41%]; 123 [0.40%]), arthralgia (198 [0.64%]; 168 [0.54%]), erythema (174 [0.56%]; 184 [0.60%]), vomiting (138 [0.44%]; 261 [0.84%]), and fever (66 [0.21%]; 124 [0.40%]) (**Table 3**).

**Table 3.** Incidence rates of adverse effects in the seven days following the date of first vaccination. For each adverse effect, incidence rates were calculated for the vaccinated (n = 31,029) and propensity matched unvaccinated (n = 30,933) cohorts as the number of positive cases divided by the total number of at risk person days during this time period. Individuals were considered at risk for developing an adverse effect from their actual or assigned date of first vaccination until they experienced the event, died, or reached the end of the 7-day study period, or until four days prior to a positive SARS-CoV-2 test. For example, in the first row, we show that 128 cases of myalgia were recorded over a total of 216,320 person-days (contributed by 31,029 vaccinated individuals), corresponding to an incidence rate of 0.59 cases per 1000 person-days.

| **Adverse effect** | **Vaccinated**<br>**Incidence Rate**<br>Cases / Person-Days<br>*[Cases Per 1000*<br>*Person-Days]* | **Unvaccinated**<br>**Incidence Rate**<br>Cases / Person-Days<br>*[Cases Per 1000*<br>*Person-Days]* | **Incidence Rate Ratio**<br>**(95% CI)** |
| --- | --- | --- | --- |
| Myalgia | 128 / 216,320 [0.59] | 123 / 215,404 [0.57] | 1 (0.8, 1.3) |
| Fever | 66 / 216,596 [0.3] | 124 / 215,447 [0.58] | 0.53 (0.39, 0.72) |
| Chills | 50 / 216,613 [0.23] | 81 / 215,564 [0.38] | 0.61 (0.42, 0.88) |
| Facial paralysis | 2 / 216,792 [0.0092] | 16 / 215,764 [0.074] | 0.12 (0.014, 0.53) |
| Erythema | 174 / 216,211 [0.8] | 184 / 215,226 [0.85] | 0.94 (0.76, 1.2) |
| Arthralgia | 198 / 216,114 [0.92] | 168 / 215,282 [0.78] | 1.2 (0.95, 1.5) |
| Vomiting | 138 / 216,328 [0.64] | 261 / 214,935 [1.2] | 0.53 (0.42, 0.65) |
| Anaphylaxis | 1 / 216,795 [0.0046] | 0 / 215,836 [0] | 0 (0.026, inf) |
| Nausea | 321 / 215,734 [1.5] | 493 / 214,132 [2.3] | 0.65 (0.56, 0.75) |
| Headache | 12 / 216,762 [0.055] | 8 / 215,829 [0.037] | 1.5 (0.56, 4.2) |
| Diarrhea | 203 / 216,173 [0.94] | 247 / 214,960 [1.1] | 0.82 (0.68, 0.99) |
| Lymphadenopathy | 78 / 216,571 [0.36] | 119 / 215,439 [0.55] | 0.65 (0.48, 0.87) |
| Fatigue | 542 / 214,896 [2.5] | 592 / 213,741 [2.8] | 0.91 (0.81, 1) |

The same events were also the most frequently documented in the 7 days following the second vaccine dose (n = 16,982 in the vaccinated cohort; n = 16,623 for the unvaccinated cohort): fatigue (200 [1.17%] vaccinated individuals; 321 [1.93%] unvaccinated individuals), nausea (143 [0.84%]; 283 [1.70%]), diarrhea (76 [0.45%]; 117 [0.70%]), myalgia (73 [0.43%]; 61 [0.37%]), arthralgia (97 [0.57%]; 91 [0.55%]), erythema (75 [0.44%]; 89 [0.54%]), vomiting (49 [0.29%]; 165 [0.99%]), and fever (31 [0.18%]; 63 [0.38%]) (**Table 4**).

**Table 4.** Incidence rates of adverse effects in the seven days following the date of second vaccination. For each adverse effect, incidence rates were calculated for the vaccinated (n = 16,982) and propensity matched unvaccinated (n = 16,623) cohorts as the number of positive cases divided by the total number of at risk person days during this time period. Individuals were considered at risk for developing an adverse effect from their actual or assigned date of first vaccination until they experienced the event, died, or reached the end of the 7-day study period, or until four days prior to a positive SARS-CoV-2 test. For example, in the first row, we show that 73 cases of myalgia were recorded over a total of 118,575 person-days (contributed by 16,982 vaccinated individuals), corresponding to an incidence rate of 0.62 cases per 1000 person-days.

| **Adverse effect** | **Vaccinated**<br>**Incidence Rate**<br>Cases / Person-Days<br>*[Cases Per 1000*<br>*Person-Days]* | **Unvaccinated**<br>**Incidence Rate**<br>Cases / Person-Days<br>*[Cases Per 1000*<br>*Person-Days]* | **Incidence Rate**<br>**Ratio (95% CI)** |
| --- | --- | --- | --- |
| Myalgia | 73 / 118,575 [0.62] | 61 / 115,041 [0.53] | 1.2 (0.82, 1.7) |
| Fever | 31 / 118,730 [0.26] | 63 / 115,024 [0.55] | 0.48 (0.3, 0.74) |
| Chills | 38 / 118,705 [0.32] | 55 / 115,026 [0.48] | 0.67 (0.43, 1) |
| Facial paralysis | 2 / 118,835 [0.017] | 2 / 115,236 [0.017] | 0.97 (0.07, 13) |
| Erythema | 75 / 118,595 [0.63] | 89 / 114,921 [0.77] | 0.82 (0.59, 1.1) |
| Arthralgia | 97 / 118,489 [0.82] | 91 / 114,915 [0.79] | 1 (0.77, 1.4) |
| Vomiting | 49 / 118,666 [0.41] | 165 / 114,627 [1.4] | 0.29 (0.2, 0.4) |
| Anaphylaxis | 0 / 118,842 [0] | 1 / 115,230 [0.0087] | 0 (0, 38) |
| Nausea | 143 / 118,331 [1.2] | 283 / 114,215 [2.5] | 0.49 (0.4, 0.6) |
| Headache | 7 / 118,814 [0.059] | 3 / 115,217 [0.026] | 2.3 (0.52, 14) |
| Diarrhea | 76 / 118,572 [0.64] | 117 / 114,888 [1] | 0.63 (0.47, 0.85) |
| Lymphadenopathy | 34 / 118,741 [0.29] | 63 / 115,005 [0.55] | 0.52 (0.33, 0.81) |
| Fatigue | 200 / 118,066 [1.7] | 321 / 113,973 [2.8] | 0.6 (0.5, 0.72) |

Notably, these rates of adverse effects documented in EHR notes were markedly lower than the rates of adverse effects observed in clinical trials and those currently captured in V-safe ^18,29,31,32^. Specifically, the rates of EHR documentation of adverse effects within 7 days of the first dose were 2.1 (95% CI: 1.5-3) to 910 (95% CI: 500-1,500) times lower than the corresponding reporting rates among the safety populations of the Phase 3 trials in the same time interval, with headache showing the largest discrepancy between these sources (**Table 5**). These differences were even more pronounced in the week following the second vaccine dose, during which EHR documentation rates were 4.9 (95% CI: 3.3-7.2) to 1,500 (95% CI: 670-2,800) times lower than corresponding reporting rates in the trials (**Table 5**). This discrepancy is to be expected, as individuals vaccinated outside of the trial or post-marketing surveillance setting are advised that it is normal to experience these adverse effects, and so they are less likely to report them to a healthcare provider. As such, the vaccine associated adverse effects which are captured in EHR notes are likely to be those that are severe or persistent enough to cause an individual to return to the clinic or otherwise notify their health care provider.

**Table 5.** Comparison of adverse effects rates in the 7 days after each vaccine dose per solicited recording in clinical trials versus unsolicited documentation in EHR notes. Clinical trial reporting rates for each adverse effect were extracted from the FDA Emergency Use Authorization documents for mRNA-1273 (Moderna) and BNT162b2 (Pfizer/BioNTech). For each cohort, the values shown are the patient counts (with percentages) who reported each symptom within 7 days of either the first or second vaccine dose. EHR documentation rates for each adverse effect were derived from Tables 3 and 4, which include the number of individuals who contributed at least one EHR note documenting the occurrence of the given symptom within 7 days of the first (Table 3) or second (Table 4) dose. To obtain the “Reporting to Documentation Ratio”, we divide the appropriate trial-derived reporting percentage by the corresponding EHR-derived documentation percentage.

| Adverse Effect | Moderna (mRNA-1273) |  | Pfizer/BioNTech (BNT162b2) |  | Mayo Clinic EHR (mRNA-1273 + BNT162b2) |  | Reporting to Documentation Ratio (Moderna / Mayo Clinic) |  | Reporting to Documentation Ratio (Pfizer / Mayo Clinic) |  |
| --- | --- | --- | --- | --- | --- | --- | --- | --- | --- | --- |
|  | Dose 1 (n = 15,168) | Dose 2 (n = 14,733) | Dose 1 (n = 4,093) | Dose 2 (n = 3,758) | Dose 1 (n = 31,609) | Dose 2 (n = 17,607) | Dose 1 [95% CI] | Dose 2 [95% CI] | Dose 1 [95% CI] | Dose 2 [95% CI] |
| Axillary swelling / tenderness / lymphadenopathy | 1553 (10.2%) | 2090 (14.2%) |  |  | 78 (0.2%) | 34 (0.2%) | 41 (33, 52) | 74 (52, 100) |  |  |
| Erythema | 430 (2.8%) | 1257 (8.6%) | 189 (4.6%) | 243 (6.5%) | 174 (0.6%) | 75 (0.4%) | 5.1 (4.3, 6.1) | 20 (16, 25) | 8.4 (6.8, 10) | 15 (12, 20) |
| Fatigue | 5635 (37.2%) | 9582 (65.3%) | 1700 (41.5%) | 2086 (55.5%) | 542 (1.7%) | 200 (1.1%) | 22 (20, 24) | 57 (50, 66) | 24 (22, 27) | 49 (42, 56) |
| Headache | 4951 (32.6%) | 8602 (58.6%) | 1413 (34.5%) | 1732 (46.1%) | 12 (0.0%) | 7 (0.0%) | 860 (470, 1400) | 1500 (670, 2800) | 910 (500, 1500) | 1200 (530, 2200) |
| Myalgia | 3441 (22.7%) | 8508 (58.0%) | 738 (18.0%) | 1260 (33.5%) | 128 (0.4%) | 73 (0.4%) | 56 (47, 66) | 140 (110, 170) | 45 (37, 53) | 81 (64, 100) |
| Arthralgia | 2511 (16.6%) | 6284 (42.8%) | 406 (9.9%) | 772 (20.5%) | 198 (0.6%) | 97 (0.6%) | 26 (23, 30) | 78 (63, 94) | 16 (13, 19) | 37 (30, 46) |
| Chills | 1253 (8.3%) | 6482 (44.2%) | 434 (10.6%) | 1114 (29.6%) | 50 (0.2%) | 38 (0.2%) | 52 (39, 68) | 200 (150, 280) | 67 (50, 89) | 140 (99, 190) |
| Nausea / Vomiting | 1262 (8.3%) | 2785 (19.0%) |  |  | 321 (1.0%) | 143 (0.8%) | 8.2 (7.3, 9.2) | 23 (20, 28) |  |  |
| Vomiting |  |  | 37 (0.9%) | 51 (1.4%) | 138 (0.4%) | 49 (0.3%) |  |  | 2.1 (1.5, 3) | 4.9 (3.3, 7.2) |
| Fever | 115 (0.8%) | 2278 (15.5%) | 222 (5.4%) | 1024 (27.2%) | 66 (0.2%) | 31 (0.2%) | 3.6 (2.7, 4.9) | 88 (61, 120) | 26 (20, 34) | 150 (110, 220) |
| Diarrhea |  |  | 402 (9.8%) | 356 (9.5%) | 203 (0.6%) | 76 (0.4%) |  |  | 15 (13, 18) | 22 (17, 28) |

### Adverse effects are not documented more frequently in the EHR notes of vaccinated individuals compared to those of propensity-matched unvaccinated individuals

To test whether documentation of each adverse effect was actually associated with receiving a COVID-19 vaccine, we computed the incidence rate ratio (IRR) for each adverse effect between the vaccinated and propensity matched unvaccinated cohorts. Several symptoms or events including headache, myalgia, arthralgia, erythema, and anaphylaxis had similar incidence rates during the 7 days after the first or second actual or assigned vaccine dose, with the confidence intervals including 1 (**Tables 3-4**). Other symptoms showed significantly lower incidence rates in the 7 days after both doses in the vaccinated cohort, including fever (IRR_dose 1_= 0.53 [0.39,-0.72]; IRR_dose 2_ = 0.48 [0.30-0.74]), nausea (IRR_dose 1_ = 0.65 [0.56-0.75]; IRR_dose 2_ = 0.49 [0.40-0.60]), vomiting (IRR_dose 1_ = 0.53 [0.42-0.65]; IRR_dose 2_ = 0.29 [0.2-0.4]), diarrhea (IRR_dose 1_ = 0.82 [0.68-0.99]; IRR_dose 2_ = 0.63 [0.47-0.85]), fatigue (IRR_dose 1_ = 0.91 [0.81-1.0]; IRR_dose 2_ = 0.6 [0.50-0.72]), chills (IRR_dose 1_ = 0.61 [0.42-0.88]; IRR_dose 2_ = 0.67 [0.43-1.0]), and lymphadenopathy (IRR_dose 1_ = 0.65 [0.48-0.87]; IRR_dose 2_ = 0.52 [0.33-0.81]).

These trends of vaccinated individuals showing similar or lower incidence rates for each considered adverse effect were also true in the 14 and 21 days following actual or assigned vaccination dates (**Tables S6-S9**). In summary, this data supports the tolerability of BNT162b2 and mRNA-1273, as individuals receiving COVID-19 vaccines do not return to the clinic to report adverse effects at higher rates than propensity matched unvaccinated individuals.

### Among matched pairs who return to clinic, adverse effects are reported at similar rates in the vaccinated and unvaccinated cohorts

To complement our previous analysis, we also considered the rates of adverse effect documentation in only the subsets of matched pairs who each contributed at least one EHR note in the 7, 14, or 21 days following the first or second actual or assigned vaccine dose. In the 7 days after the first actual or assigned vaccination date (n = 1,399 propensity matched pairs) the IRR 95% CIs for most adverse effects included 1 (**Table 6**). Only nausea, vomiting and diarrhea showed differences in incidence rates, with lower incidence rates in the vaccinated cohort (IRR_nausea_ = 0.59 [0.41-0.83]; IRR_diarrhea_ = 0.50 [0.30-0.80]; IRR_vomiting_ = 0.59 [0.37-0.94]). (**Table 6**). After the second vaccination date (n = 897 propensity matched pairs), myalgia was documented more frequently among vaccinated individuals (IRR_myalgia_ = 2.5 [1.1-6.7]), while all other adverse effects were recorded at similar or lower rates in vaccinated compared to unvaccinated individuals (**Table 7**).

**Table 6.** Incidence rates of adverse effects in the seven days following the date of first vaccination for all individuals with at least one clinical note recorded during this time. For each adverse effect, incidence rates were calculated for the vaccinated and propensity matched unvaccinated cohorts (n = 1,399 pairs) as the number of positive cases divided by the total number of at risk person days during this time period. Individuals were considered at risk for developing an adverse effect from their actual or assigned date of first vaccination until they experienced the event, died, or reached the end of the 7-day study period, or until four days prior to a positive SARS-CoV-2 test. For example, in the first row, we show that 31 cases of myalgia were recorded over a total of 9,623 person-days (contributed by 1,399 vaccinated individuals), corresponding to an incidence rate of 3.2 cases per 1000 person-days.

| **Adverse effect** | **Vaccinated**<br>**Incidence Rate**<br>Cases / Person-Days<br>*[Cases Per 1000*<br>*Person-Days]* | **Unvaccinated**<br>**Incidence Rate**<br>Cases / Person-Days<br>*[Cases Per 1000*<br>*Person-Days]* | **Incidence Rate**<br>**Ratio (95% CI)** |
| --- | --- | --- | --- |
| Myalgia | 31 / 9,623 [3.2] | 27 / 9,634 [2.8] | 1.1 (0.66, 2) |
| Fever | 19 / 9,698 [2] | 24 / 9,663 [2.5] | 0.79 (0.41, 1.5) |
| Chills | 9 / 9,711 [0.93] | 9 / 9,705 [0.93] | 1 (0.35, 2.8) |
| Facial paralysis | 0 / 9,752 [0] | 2 / 9,731 [0.21] | 0 (0, 5.3) |
| Erythema | 36 / 9,613 [3.7] | 49 / 9,576 [5.1] | 0.73 (0.46, 1.1) |
| Arthralgia | 49 / 9,583 [5.1] | 41 / 9,577 [4.3] | 1.2 (0.77, 1.9) |
| Vomiting | 31 / 9,649 [3.2] | 52 / 9,555 [5.4] | 0.59 (0.37, 0.94) |
| Anaphylaxis | 0 / 9,752 [0] | 0 / 9,738 [0] | NA |
| Nausea | 55 / 9,575 [5.7] | 92 / 9,429 [9.8] | 0.59 (0.41, 0.83) |
| Headache | 1 / 9,752 [0.1] | 4 / 9,733 [0.41] | 0.25 (0.0051, 2.5) |
| Diarrhea | 28 / 9,645 [2.9] | 56 / 9,591 [5.8] | 0.5 (0.3, 0.8) |
| Lymphadenopathy | 18 / 9,687 [1.9] | 29 / 9,637 [3] | 0.62 (0.32, 1.2) |
| Fatigue | 101 / 9,367 [11] | 122 / 9,311 [13] | 0.82 (0.63, 1.1) |

**Table 7.** Incidence rates of adverse effects in the seven days following the date of second vaccination for all individuals with at least one clinical note recorded during this time. For each adverse effect, incidence rates were calculated for the vaccinated and propensity matched unvaccinated cohorts (n = 897 pairs) as the number of positive cases divided by the total number of at risk person days during this time period. Individuals were considered at risk for developing an adverse effect from their actual or assigned date of first vaccination until they experienced the event, died, or reached the end of the 7-day study period, or until four days prior to a positive SARS-CoV-2 test. For example, in the first row, we show that 21 cases of myalgia were recorded over a total of 6,191 person-days (contributed by 897 vaccinated individuals), corresponding to an incidence rate of 3.4 cases per 1000 person-days.

| **Adverse effect** | **Vaccinated**<br>**Incidence Rate**<br>Cases / Person-Days<br>*[Cases Per 1000*<br>*Person-Days]* | **Unvaccinated**<br>**Incidence Rate**<br>Cases / Person-Days<br>*[Cases Per 1000*<br>*Person-Days]* | **Incidence Rate Ratio**<br>**(95% CI)** |
| --- | --- | --- | --- |
| Myalgia | 21 / 6,191 [3.4] | 8 / 6,008 [1.3] | 2.5 (1.1, 6.7) |
| Fever | 9 / 6,238 [1.4] | 14 / 5,978 [2.3] | 0.62 (0.24, 1.5) |
| Chills | 8 / 6,238 [1.3] | 14 / 5,971 [2.3] | 0.55 (0.2, 1.4) |
| Facial paralysis | 1 / 6,271 [0.16] | 0 / 6,022 [0] | 0 (0.025, inf) |
| Erythema | 13 / 6,220 [2.1] | 17 / 5,959 [2.9] | 0.73 (0.33, 1.6) |
| Arthralgia | 22 / 6,210 [3.5] | 20 / 5,956 [3.4] | 1.1 (0.55, 2) |
| Vomiting | 8 / 6,241 [1.3] | 32 / 5,912 [5.4] | 0.24 (0.094, 0.52) |
| Anaphylaxis | 0 / 6,273 [0] | 1 / 6,015 [0.17] | 0 (0, 37) |
| Nausea | 29 / 6,158 [4.7] | 50 / 5,837 [8.6] | 0.55 (0.34, 0.89) |
| Headache | 2 / 6,266 [0.32] | 0 / 6,022 [0] | 0 (0.18, inf) |
| Diarrhea | 19 / 6,200 [3.1] | 22 / 5,957 [3.7] | 0.83 (0.42, 1.6) |
| Lymphadenopathy | 6 / 6,250 [0.96] | 13 / 5,970 [2.2] | 0.44 (0.14, 1.2) |
| Fatigue | 52 / 6,075 [8.6] | 64 / 5,764 [11] | 0.77 (0.52, 1.1) |

Similarly, vaccinated individuals who returned to the clinic also showed comparable or lower incidence rates for each considered adverse effect in the 14 and 21 days post vaccination compared to their matched unvaccinated individuals who also returned to the clinic (**Tables S10-S13**). Taken together, this analysis further corroborates the conclusion that most adverse effects, with the potential exception of myalgia in the week following second vaccination, are not documented in the EHRs of vaccinated individuals at unexpectedly high rates.

## Discussion

This study demonstrates that the two currently FDA-authorized COVID-19 vaccines, BNT162b2 and mRNA-1273, are safe and tolerated beyond the confines of a clinical trial setting. This conclusion is consistent with the extensive safety and tolerability assessments conducted in Phase 1/2 and Phase 3 trials over the past nine months and in the post-marketing surveillance efforts which are currently under way^4,5,18,33,34^. Here we assessed real world safety and tolerability by longitudinally curating the EHR documentation of adverse effects in 31,029 individuals receiving at least one dose of a COVID-19 vaccine compared to a propensity matched unvaccinated cohort of similar size. Compared to this control cohort, vaccinated individuals were not more likely to return to the clinic (including the emergency department) or to report any of the surveyed adverse effects within 7, 14, or 21 days after the first or second vaccine dose. When considering only the subsets of matched pairs who each had at least one clinical note in these defined time intervals, myalgia was the only symptom with a modest increase in documentation frequency among vaccinated individuals, while all other potential side effects occurred at similar or lower rates in this cohort.

The purpose of using propensity matching in this study was to establish an expected frequency for each potential vaccine associated adverse effect in a group of individuals with similar demographic and geographic characteristics, akin to a placebo group in a randomized clinical trial. Our finding that EHR notes from vaccinated and propensity matched unvaccinated individuals record similar rates of potential vaccine associated adverse effects differs from the data obtained in Phase 3 trials, wherein vaccinated participants experienced higher rates of symptoms than those receiving placebo. Further, the absolute rates of adverse effects documented in these EHR notes are well below the rates reported in clinical trials (see **Table 5**) and in the active post-marketing surveillance of these vaccines (V-safe) (**Table S14**). These discrepancies are likely attributable to differences in the populations analyzed and the methodology of reporting symptoms.

Regarding the populations analyzed, it is important to realize that adverse effects experienced by healthcare workers, who comprise a significant proportion of the Phase 1a vaccinated population thus far, are generally less likely to be documented in EHR notes. At many institutions, healthcare workers experiencing potential vaccine side effects are directed to follow up with Occupational Health Services, which is an entity of the employer and thus does not document the reported symptoms in EHR notes. Several institutions have also established COVID-19 response lines, which will usually direct individuals experiencing nonspecific vaccine side effects toward self-care. Further, healthcare workers may be less likely to report adverse effects at baseline given their clinical expertise and ability to self-assess the severity of their illness.

Regarding reporting methods, both trials included a 7-day post-vaccination period in which symptoms were actively solicited from some or all individuals as well as longer periods in which unsolicited adverse effects and serious adverse reactions were recorded from all individuals. V-safe similarly solicits adverse effect reports from its voluntary participants on a regular basis. In contrast, our methods rely exclusively on the recording of unsolicited symptoms or events in the EHR. Given that individuals are warned of the likely vaccine associated adverse effects at the time of vaccination in the real world setting, it is likely that most mild or moderate symptoms are never actually reported and thus are not documented in an EHR note.

That said, serious safety concerns by definition require medical care and thus are likely to be documented in the EHR. For example, should an individual experience anaphylaxis, this individual will likely require emergent care, during which one or more clinical notes will be written and will mention this phenotype. Thus, our method should identify the symptoms and phenotypes that represent the most serious threats to vaccine safety and tolerability of practical significance. Indeed, this is the central reason why our analysis should be viewed as complementary to the data which has been obtained in the more controlled setting of clinical trials. It is extremely important to know and understand the frequencies of side effects which were observed via solicitation in the trial setting, and this study does not contradict or refute this data in any way. Rather, our assessment specifically aims to describe the frequencies of adverse effects that receive some form of clinical attention as evidenced by their documentation in the EHR.

In light of this, it is worth noting that the rate of severe side effects was actually similar among participants receiving vaccine or placebo in the Pfizer/BioNTech and Moderna Phase 3 trials at 2% or less, with the exception of severe fatigue in 3.8% of participants after their second dose of BNT162b2^4,5^. This is consistent with our observations that vaccinated and unvaccinated individuals have similar EHR documentation rates for each surveyed potential vaccine associated symptom. And our finding that vaccinated individuals are not more likely to receive emergent care provides further orthogonal support for the safety of these vaccines.

There are several limitations of this study. First, while the analysis was conducted on a population derived from a large healthcare system, the cohort demographics are not representative of the American population. For example, both the vaccinated and unvaccinated cohorts were predominantly Caucasian (>90%) and female (>60%). These biases likely reflect both the populations who receive care at the various Mayo Clinic centers and the populations who have been prioritized in Phase 1a of the vaccine rollout. The enrichment of healthcare workers among individuals vaccinated in this phase likely leads to an underestimation of the rates of return to clinic, due to the factors previously discussed (i.e. access to an Occupational Health Services office and other institution-specific COVID-19 response centers). However, it is worth noting that even those individuals who utilize such services are likely to generate EHR notes if they are indeed determined to require clinical attention or intervention. Second, the BERT model used to curate EHR notes does not imply a direct link between COVID-19 vaccination and the experience of a phenotype. That is, we simply capture the occurrence of an adverse effect without ensuring that the clinical note indeed suggests or confirms that vaccination caused the symptom. This shortcoming is addressed by comparing vaccinated individuals to the unvaccinated control cohort, which establishes a baseline expected frequency for each symptom in the absence of vaccination. Finally, while sentences suggesting the occurrence of anaphylaxis were manually reviewed to confirm both the positive sentiment and the tense (see **Methods**), sentences for the other curated phenotypes were not reviewed. In the future, we will train natural language processing models to discriminate past from present tense, thereby circumventing the need for this manual review.

As the remainder of the US population undergoes COVID-19 vaccination, it will not be feasible to conduct solicited reporting of adverse effects in all vaccinated individuals. Augmented curation of EHR notes for real world safety monitoring presents a practical solution to this problem which can be deployed to complement other directed surveillance efforts like V-safe. The method demonstrated here represents a scalable approach to continuously monitor serious safety concerns associated with any authorized COVID-19 vaccines. Taken together with our recent study highlighting the real world efficacy of these vaccines^24^, this data reinforces that individuals, providers, and public health officials should proceed rapidly with vaccination efforts, with high confidence in their efficacy and safety.

## Data Availability

After publication, the data will be made available upon reasonable requests to the corresponding author. A proposal with detailed description of study objectives and the statistical analysis plan will be needed for evaluation of the reasonability of requests. Deidentified data will be provided after approval from the corresponding author and the Mayo Clinic.

## Acknowledgements

The authors thank Murali Aravamudan for the careful review and feedback on this manuscript.

## Author Contributions

VS, PL, and SA conceived the study. RM, PL, AJV and VS wrote the manuscript and reviewed the findings. ES, AP, SA, CP, VA, PA, AR, CC, KC, DD, NK, ER, GB, AM, TW contributed methods, analysis, and software. JCOH, GJG, AWW, ADB, MDS, AV, and JH reviewed the study, findings, and the manuscript. All authors revised the manuscript.

## Supplemental Material

**Table S1.** Number of matched pairs in which both the vaccinated individual and their propensity matched unvaccinated individual had at least one clinical note in the specified time intervals. The first column gives the time interval: within 7, 14, or 21 days. The second column gives the number of matched pairs with at least one clinical note in the defined time interval after the first actual or assigned vaccination date. The third column gives the number of matched pairs with at least one clinical note in the defined time interval after the second actual or assigned vaccination date.

| Time Interval | Matched Pairs With At Least One EHR Note After First Dose | Matched Pairs With At Least One EHR Note After Second Dose |
| --- | --- | --- |
| Within 7 Days | 1,399 | 897 |
| Within 14 Days | 2,643 | 1,717 |
| Within 21 Days | 3,553 | 2,360 |

**Table S2.** Number of Emergency Department (ED) notes contributed by vaccinated and unvaccinated individuals during the 7, 14, and 21 days after the first actual or assigned vaccination date. The percentages shown were obtained by dividing the number of ED notes by the total number of clinical notes for the cohort in the given time interval, and multiplying by 100. To assess the magnitude and significance of difference between the percentages of ED notes, the odds ratio (OR) and corresponding 95% CI are shown. With the null hypothesis that the OR falls between 0.91 and 1.1, a difference was considered significant if the upper bound of 95% CI was less than 0.91 or the lower bound of the 95% CI was greater than 1.1.

| **Time Interval After**<br>**First Actual or**<br>**Assigned Dose** | **Vaccinated Cohort**<br>**(n = 31,029)**<br>Number of ED Notes<br>(% of All Notes) | **Unvaccinated Cohort**<br>**(n = 30,933)**<br>Number of ED Notes<br>(% of All Notes) | **Odds Ratio**<br>**(95% CI)** |
| --- | --- | --- | --- |
| **1 Day** | 107<br>(3.00%) | 182<br>(3.21%) | 0.93<br>(0.72 - 1.20) |
| **7 Days** | 496<br>(3.74%) | 812<br>(3.98%) | 0.94<br>(0.83 - 1.05) |
| **14 Days** | 794<br>(3.42%) | 1355<br>(3.86%) | 0.88<br>(0.81 - 0.97) |
| **21 Days** | 1009<br>(3.25%) | 1866<br>(3.89%) | 0.83<br>(0.77 - 0.90) |

**Table S3.** Number of Emergency Department (ED) notes contributed by vaccinated and unvaccinated individuals (n = 31,069 each) during the 7, 14, and 21 days after the second actual or assigned vaccination date. The percentages shown were obtained by dividing the number of ED notes by the total number of clinical notes for the cohort in the given time interval, and multiplying by 100. To assess the magnitude and significance of difference between the percentages of ED notes, the odds ratio (OR) and corresponding 95% CI are shown. With the null hypothesis that the OR falls between 0.91 and 1.1, a difference was considered significant if the upper bound of 95% CI was less than 0.91 or the lower bound of the 95% CI was greater than 1.1.

| **Time Interval After**<br>**Second Actual or**<br>**Assigned Dose** | **Vaccinated Cohort**<br>**(n = 16,982 individuals)**<br>Number of ED Notes<br>(% of All Notes) | **Unvaccinated Cohort**<br>**(n = 16,623)**<br>Number of ED Notes<br>(% of All Notes) | **Odds Ratio**<br>**(95% CI)** |
| --- | --- | --- | --- |
| **1 Day** | 49<br>(3.40%) | 136<br>(4.47%) | 0.75<br>(0.53 - 1.06)) |
| **7 Days** | 152<br>(3.39%) | 368<br>(3.87%) | 0.87<br>(0.71,1.06) |
| **14 Days** | 208<br>(2.92%) | 532<br>(3.65%) | 0.79<br>(0.67 - 0.94) |
| **21 Days** | 253<br>(2.98%) | 684<br>(3.93%) | 0.75<br>(0.64 - 0.87) |

**Table S4.** Number of individuals contributing Emergency Department (ED) notes in the vaccinated and unvaccinated cohorts during the 1, 7, 14, and 21 days after the first actual or assigned vaccination date. Percentages shown are obtained by dividing the number of individuals with at least one ED note by the total number of individuals in the cohort, and multiplying by 100. To assess the magnitude and significance of difference between the percentages of patients with ED notes, the odds ratio (OR) and corresponding 95% CI are shown. With the null hypothesis that the OR falls between 0.91 and 1.1, a difference was considered significant if the upper bound of 95% CI was less than 0.91 or the lower bound of the 95% CI was greater than 1.1.

| **Time Interval**<br>**After First**<br>**Actual or**<br>**Assigned Dose** | **Vaccinated Cohort**<br>**(n = 31,029)**<br>Individuals with ED Visits<br>(% of Cohort) | **Unvaccinated Cohort**<br>**(n = 30,933)**<br>Individuals with ED Visits<br>(% of Cohort) | **Odds Ratio**<br>**(95% CI)** |
| --- | --- | --- | --- |
| **1 Day** | 87<br>(0.28%) | 151<br>(0.49%) | 0.57<br>(0.43 - 0.75) |
| **7 Days** | 376<br>(1.21%) | 623<br>(2.01%) | 0.60<br>(0.52 - 0.68) |
| **14 Days** | 592<br>(1.91%) | 1,003<br>(3.24%) | 0.58<br>(0.52 - 0.64) |
| **21 Days** | 753<br>(2.43%) | 1,352<br>(4.37%) | 0.54<br>(0.50 - 0.60) |

**Table S5.** Number of individuals contributing Emergency Department (ED) notes in the vaccinated and unvaccinated cohorts during the 1, 7, 14, and 21 days after the second actual or assigned vaccination date. Percentages shown are obtained by dividing the number of individuals with at least one ED note by the total number of individuals in the cohort, and multiplying by 100. To assess the magnitude and significance of difference between the percentages of patients with ED notes, the odds ratio (OR) and corresponding 95% CI are shown. With the null hypothesis that the OR falls between 0.91 and 1.1, a difference was considered significant if the upper bound of 95% CI was less than 0.91 or the lower bound of the 95% CI was greater than 1.1.

| **Time Interval**<br>**After Second**<br>**Actual or**<br>**Assigned Dose** | **Vaccinated Cohort**<br>**(n = 16,982)**<br>Individuals with ED Visits<br>(% of Cohort) | **Unvaccinated Cohort**<br>**(n = 16,623)**<br>Individuals with ED Visits<br>(% of Cohort) | **Odds Ratio**<br>**(95% CI)** |
| --- | --- | --- | --- |
| **1 Day** | 39<br>(0.23%) | 104<br>(0.63%) | 0.37<br>(0.25 - 0.53) |
| **7 Days** | 122<br>(0.72%) | 284<br>(1.71%) | 0.42<br>(0.33 - 0.52) |
| **14 Days** | 164<br>(0.97%) | 407<br>(2.45%) | 0.39<br>(0.32 - 0.47) |
| **21 Days** | 200<br>(1.18%) | 508<br>(3.06%) | 0.38<br>(0.32 - 0.45) |

**Table S6.** Incidence rates of adverse effects in the 14 days following the date of first vaccination. For each adverse effect, incidence rates were calculated for the vaccinated (n = 31,029) and propensity matched unvaccinated (n = 30,933) cohorts as the number of positive cases divided by the total number of at risk person days during this time period. Individuals were considered at risk for developing an adverse effect from their actual or assigned date of first vaccination until they experienced the event, died, or reached the end of the 14-day study period, or until four days prior to a positive SARS-CoV-2 test. For example, in the first row, we show that 164 cases of myalgia were recorded over a total of 431,530 person-days (contributed by 31,029 vaccinated individuals), corresponding to an incidence rate of 0.38 cases per 1000 person-days.

| **Adverse effect** | **Vaccinated**<br>**Incidence Rate**<br>Cases / Person-Days<br>*[Cases Per 1000*<br>*Person-Days]* | **Unvaccinated**<br>**Incidence Rate**<br>Cases/Person-Days<br>*[Cases Per 1000*<br>*Person-Days]* | **Incidence Rate**<br>**Ratio (95% CI)** |
| --- | --- | --- | --- |
| Myalgia | 164 / 431,530 [0.38] | 171 / 428,939 [0.4] | 0.95 (0.76, 1.2) |
| Fever | 94 / 432,266 [0.22] | 172 / 429,113 [0.4] | 0.54 (0.42, 0.7) |
| Chills | 66 / 432,413 [0.15] | 115 / 429,357 [0.27] | 0.57 (0.41, 0.78) |
| Facial paralysis | 6 / 432,925 [0.014] | 17 / 430,117 [0.04] | 0.35 (0.11, 0.93) |
| Erythema | 220 / 431,207 [0.51] | 262 / 428,308 [0.61] | 0.83 (0.69, 1) |
| Arthralgia | 237 / 430,887 [0.55] | 221 / 428,531 [0.52] | 1.1 (0.88, 1.3) |
| Vomiting | 177 / 431,536 [0.41] | 333 / 427,684 [0.78] | 0.53 (0.44, 0.63) |
| Anaphylaxis | 1 / 432,940 [0.0023] | 0 / 430,298 [0] | 0 (0.025, inf) |
| Nausea | 366 / 429,968 [0.85] | 613 / 425,285 [1.4] | 0.59 (0.52, 0.67) |
| Headache | 12 / 432,830 [0.028] | 10 / 430,229 [0.023] | 1.2 (0.47, 3.1) |
| Diarrhea | 238 / 431,101 [0.55] | 328 / 427,689 [0.77] | 0.72 (0.61, 0.85) |
| Lymphadenopathy | 93 / 432,225 [0.22] | 153 / 429,049 [0.36] | 0.6 (0.46, 0.79) |
| Fatigue | 596 / 427,727 [1.4] | 745 / 424,096 [1.8] | 0.79 (0.71, 0.88) |

**Table S7.**
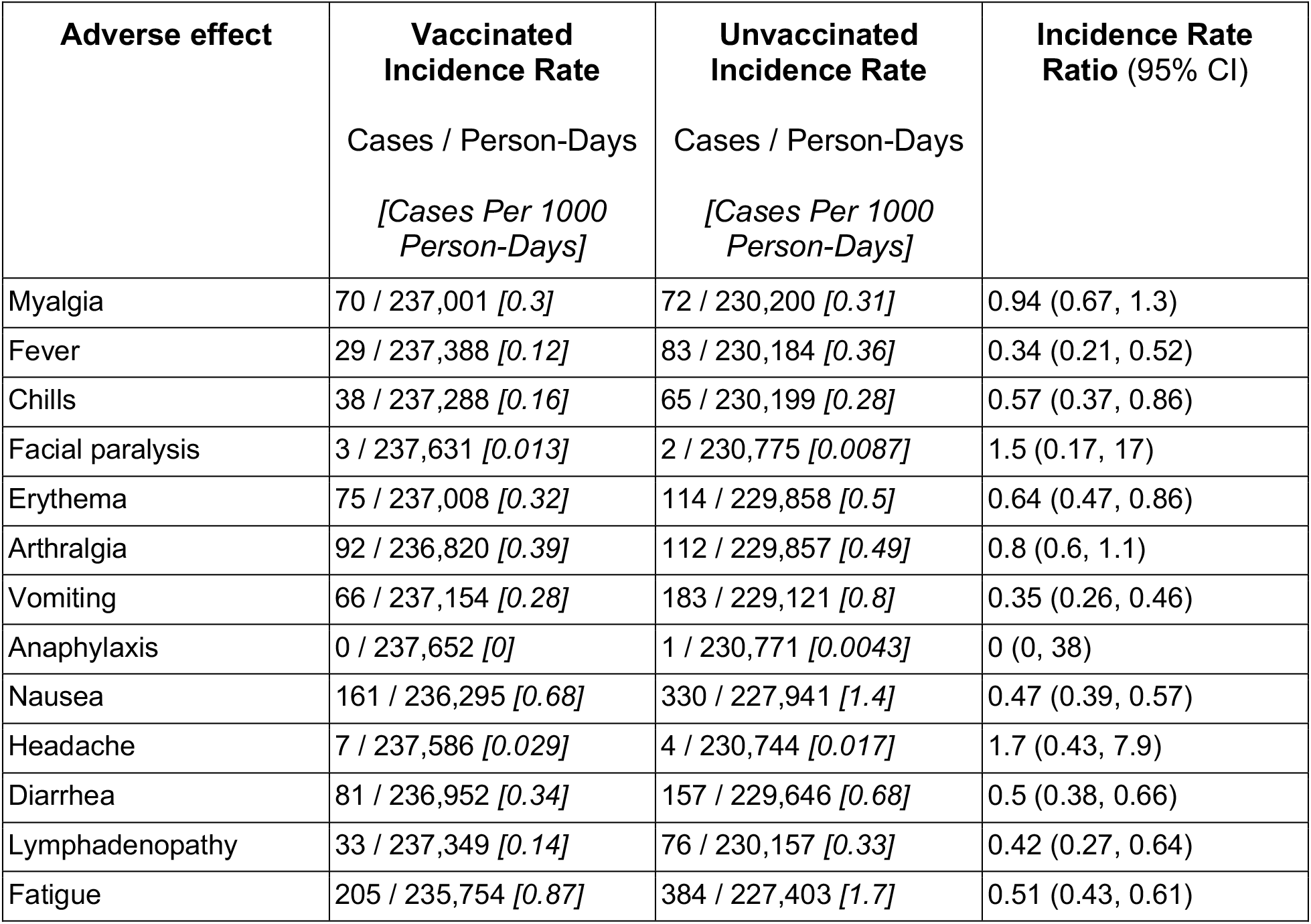
Incidence rates of adverse effects in the 14 days following the date of second vaccination. For each adverse effect, incidence rates were calculated for the vaccinated (n = 16,982) and propensity matched unvaccinated (n = 16,623) cohorts as the number of positive cases divided by the total number of at risk person days during this time period. Individuals were considered at risk for developing an adverse effect from their actual or assigned date of first vaccination until they experienced the event, died, or reached the end of the 14-day study period, or until four days prior to a positive SARS-CoV-2 test. For example, in the first row, we show that 70 cases of myalgia were recorded over a total of 237,001 person-days (contributed by 16,982 vaccinated individuals), corresponding to an incidence rate of 0.3 cases per 1000 person-days.

**Table S8.** Incidence rates of adverse effects in the 21 days following the date of first vaccination. For each adverse effect, incidence rates were calculated for the vaccinated (n = 31,029) and propensity matched unvaccinated (n = 30,933) cohorts as the number of positive cases divided by the total number of at risk person days during this time period. Individuals were considered at risk for developing an adverse effect from their actual or assigned date of first vaccination until they experienced the event, died, or reached the end of the 21-day study period, or until four days prior to a positive SARS-CoV-2 test. For example, in the first row, we show that 255 cases of myalgia were recorded over a total of 645,745 person-days (contributed by 31,029 vaccinated individuals), corresponding to an incidence rate of 0.39cases per 1000 person-days.

| **Adverse effect** | **Vaccinated**<br>**Incidence Rate**<br>Cases / Person-Days<br>*[Cases Per 1000*<br>*Person-Days]* | **Unvaccinated**<br>**Incidence Rate**<br>Cases / Person-Days<br>*[Cases Per 1000*<br>*Person-Days]* | **Incidence Rate**<br>**Ratio (95% CI)** |
| --- | --- | --- | --- |
| Myalgia | 255 / 645,745 [0.39] | 261 / 640,960 [0.41] | 0.97 (0.81, 1.2) |
| Fever | 131 / 647,335 [0.2] | 242 / 641,335 [0.38] | 0.54 (0.43, 0.67) |
| Chills | 103 / 647,642 [0.16] | 177 / 641,844 [0.28] | 0.58 (0.45, 0.74) |
| Facial paralysis | 8 / 648,723 [0.012] | 24 / 643,566 [0.037] | 0.33 (0.13, 0.76) |
| Erythema | 333 / 644,910 [0.52] | 388 / 639,664 [0.61] | 0.85 (0.73, 0.99) |
| Arthralgia | 359 / 644,408 [0.56] | 310 / 640,157 [0.48] | 1.2 (0.99, 1.3) |
| Vomiting | 281 / 645,610 [0.44] | 482 / 638,286 [0.76] | 0.58 (0.5, 0.67) |
| Anaphylaxis | 1 / 648,790 [0.0015] | 1 / 643,874 [0.0016] | 0.99 (0.013, 78) |
| Nausea | 590 / 642,070 [0.92] | 894 / 633,475 [1.4] | 0.65 (0.59, 0.72) |
| Headache | 17 / 648,596 [0.026] | 12 / 643,729 [0.019] | 1.4 (0.63, 3.2) |
| Diarrhea | 355 / 644,837 [0.55] | 477 / 638,573 [0.75] | 0.74 (0.64, 0.85) |
| Lymphadenopathy | 152 / 647,178 [0.23] | 212 / 641,412 [0.33] | 0.71 (0.57, 0.88) |
| Fatigue | 905 / 637,734 [1.4] | 1103 / 630,971 [1.7] | 0.81 (0.74, 0.89) |

**Table S9.** Incidence rates of adverse effects in the 21 days following the date of second vaccination. For each adverse effect, incidence rates were calculated for the vaccinated (n = 16,982) and propensity matched unvaccinated (n = 16,623) cohorts as the number of positive cases divided by the total number of at risk person days during this time period. Individuals were considered at risk for developing an adverse effect from their actual or assigned date of first vaccination until they experienced the event, died, or reached the end of the 21-day study period, or until four days prior to a positive SARS-CoV-2 test. For example, in the first row, we show that 98 cases of fatigue were recorded over a total of 355,085 person-days (contributed by 16,982 vaccinated individuals), corresponding to an incidence rate of 0.28 cases per 1000 person-days.

| **Adverse effect** | **Vaccinated**<br>**Incidence Rate**<br>Cases / Person-Days<br>*[Cases Per 1000*<br>*Person-Days]* | **Unvaccinated**<br>**Incidence Rate**<br>Cases / Person-Days<br>*[Cases Per 1000*<br>*Person-Days]* | **Incidence Rate**<br>**Ratio (95% CI)** |
| --- | --- | --- | --- |
| Myalgia | 98 / 355,085 [0.28] | 94 / 344,935 [0.27] | 1 (0.76, 1.4) |
| Fever | 38 / 355,930 [0.11] | 114 / 344,812 [0.33] | 0.32 (0.22, 0.47) |
| Chills | 49 / 355,724 [0.14] | 89 / 344,912 [0.26] | 0.53 (0.37, 0.76) |
| Facial paralysis | 3 / 356,402 [0.0084] | 3 / 346,104 [0.0087] | 0.97 (0.13, 7.3) |
| Erythema | 100 / 355,102 [0.28] | 150 / 344,240 [0.44] | 0.65 (0.5, 0.84) |
| Arthralgia | 129 / 354,788 [0.36] | 151 / 344,213 [0.44] | 0.83 (0.65, 1.1) |
| Vomiting | 97 / 355,258 [0.27] | 248 / 342,827 [0.72] | 0.38 (0.3, 0.48) |
| Anaphylaxis | 0 / 356,444 [0] | 1 / 346,113 [0.0029] | 0 (0, 38) |
| Nausea | 221 / 353,532 [0.63] | 447 / 340,308 [1.3] | 0.48 (0.4, 0.56) |
| Headache | 10 / 356,301 [0.028] | 5 / 346,062 [0.014] | 1.9 (0.6, 7.2) |
| Diarrhea | 112 / 354,940 [0.32] | 220 / 343,646 [0.64] | 0.49 (0.39, 0.62) |
| Lymphadenopathy | 50 / 355,769 [0.14] | 97 / 344,888 [0.28] | 0.5 (0.35, 0.71) |
| Fatigue | 287 / 352,482 [0.81] | 501 / 339,423 [1.5] | 0.55 (0.48, 0.64) |

**Table S10.** Incidence rates of adverse effects in the 14 days following the date of first vaccination for all individuals with at least one clinical note recorded during this time. For each adverse effect, incidence rates were calculated for the propensity matched vaccinated and unvaccinated individuals (n = 2,643 pairs) as the number of positive cases divided by the total number of at risk person days during this time period. Individuals were considered at risk for developing an adverse effect from their actual or assigned date of first vaccination until they experienced the event, died, or reached the end of the 14-day study period, or until four days prior to a positive SARS-CoV-2 test. For example, in the first row, we show that 65 cases of myalgia were recorded over a total of 36,105 person-days (contributed by 2,643 vaccinated individuals), corresponding to an incidence rate of 1.8 cases per 1000 person-days.

| **Adverse effect** | **Vaccinated**<br>**Incidence Rate**<br>Cases / Person-Days<br>*[Cases Per 1000*<br>*Person-Days]* | **Unvaccinated**<br>**Incidence Rate**<br>Cases / Person-Days<br>*[Cases Per 1000*<br>*Person-Days]* | **Incidence Rate**<br>**Ratio (95% CI)** |
| --- | --- | --- | --- |
| Myalgia | 65 / 36,105 [1.8] | 68 / 35,877 [1.9] | 0.95 (0.67, 1.4) |
| Fever | 38 / 36,394 [1] | 65 / 35,965 [1.8] | 0.58 (0.38, 0.88) |
| Chills | 17 / 36,485 [0.47] | 40 / 36,094 [1.1] | 0.42 (0.22, 0.76) |
| Facial paralysis | 3 / 36,638 [0.082] | 4 / 36,315 [0.11] | 0.74 (0.11, 4.4) |
| Erythema | 79 / 36,098 [2.2] | 95 / 35,688 [2.7] | 0.82 (0.6, 1.1) |
| Arthralgia | 106 / 35,928 [3] | 84 / 35,779 [2.3] | 1.3 (0.93, 1.7) |
| Vomiting | 65 / 36,179 [1.8] | 119 / 35,539 [3.3] | 0.54 (0.39, 0.73) |
| Anaphylaxis | 1 / 36,634 [0.027] | 0 / 36,359 [0] | 0 (0.025, inf) |
| Nausea | 139 / 35,714 [3.9] | 216 / 34,904 [6.2] | 0.63 (0.5, 0.78) |
| Headache | 6 / 36,610 [0.16] | 5 / 36,322 [0.14] | 1.2 (0.3, 4.9) |
| Diarrhea | 65 / 36,207 [1.8] | 122 / 35,569 [3.4] | 0.52 (0.38, 0.71) |
| Lymphadenopathy | 44 / 36,369 [1.2] | 67 / 35,946 [1.9] | 0.65 (0.43, 0.96) |
| Fatigue | 226 / 34,960 [6.5] | 296 / 34,290 [8.6] | 0.75 (0.63, 0.89) |

**Table S11.**
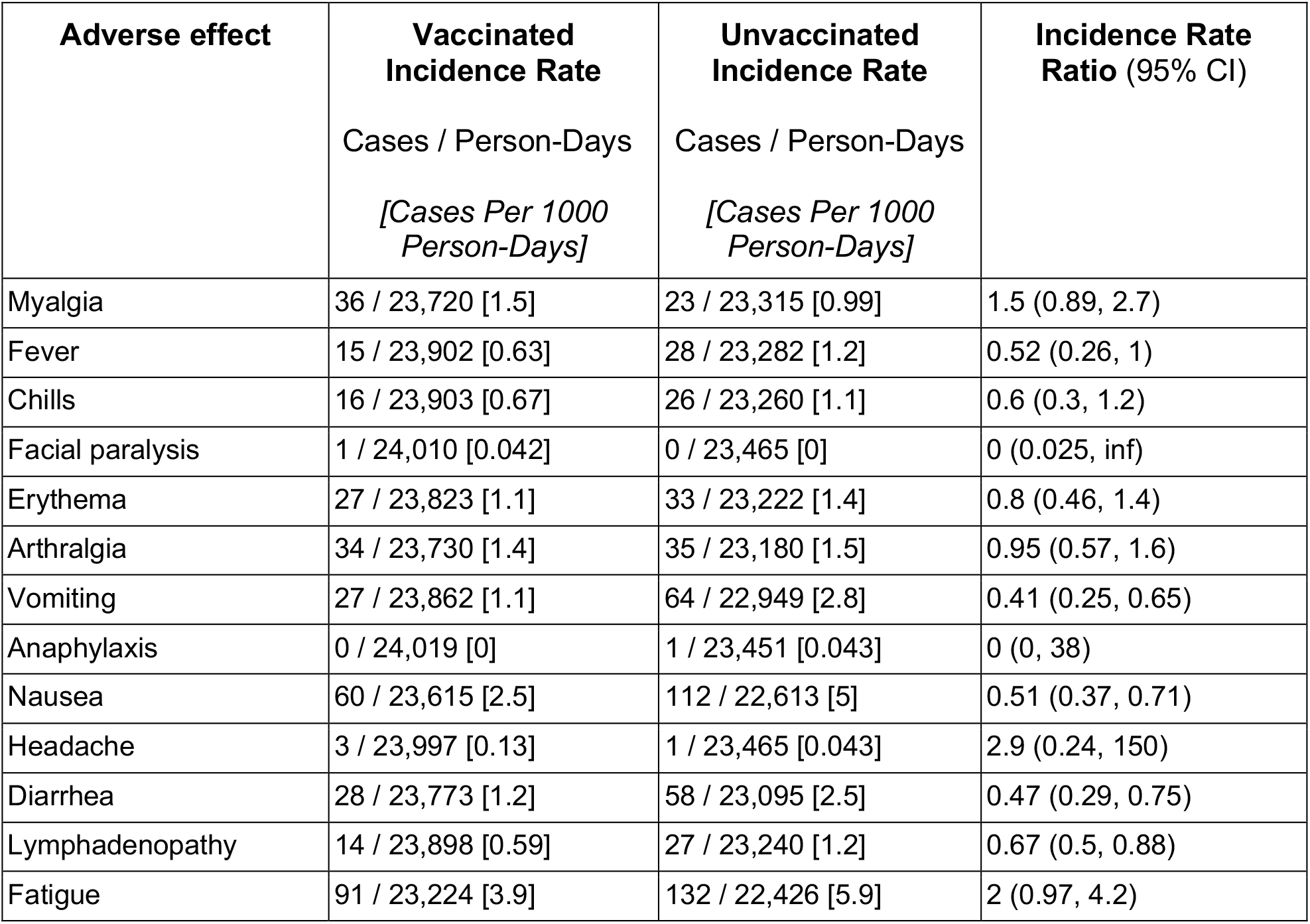
Incidence rates of adverse effects in the 14 days following the date of second vaccination for all individuals with at least one clinical note recorded during this time. For each adverse effect, incidence rates were calculated for the propensity matched vaccinated and unvaccinated individuals (n = 1,717 pairs) as the number of positive cases divided by the total number of at risk person days during this time period. Individuals were considered at risk for developing an adverse effect from their actual or assigned date of first vaccination until they experienced the event, died, or reached the end of the 14-day study period, or until four days prior to a positive SARS-CoV-2 test. For example, in the first row, we show that 36 cases of myalgia were recorded over a total of 23,720 person-days (contributed by 1,717 vaccinated individuals), corresponding to an incidence rate of 1.5 cases per 1000 person-days.

| **Adverse effect** | **Vaccinated**<br>**Incidence Rate**<br>Cases / Person-Days<br>*[Cases Per 1000*<br>*Person-Days]* | **Unvaccinated**<br>**Incidence Rate**<br>Cases / Person-Days<br>*[Cases Per 1000*<br>*Person-Days]* | **Incidence Rate**<br>**Ratio (95% CI)** |
| --- | --- | --- | --- |
| Myalgia | 36 / 23,720 [1.5] | 23 / 23,315 [0.99] | 1.5 (0.89, 2.7) |
| Fever | 15 / 23,902 [0.63] | 28 / 23,282 [1.2] | 0.52 (0.26, 1) |
| Chills | 16 / 23,903 [0.67] | 26 / 23,260 [1.1] | 0.6 (0.3, 1.2) |
| Facial paralysis | 1 / 24,010 [0.042] | 0 / 23,465 [0] | 0 (0.025, inf) |
| Erythema | 27 / 23,823 [1.1] | 33 / 23,222 [1.4] | 0.8 (0.46, 1.4) |
| Arthralgia | 34 / 23,730 [1.4] | 35 / 23,180 [1.5] | 0.95 (0.57, 1.6) |
| Vomiting | 27 / 23,862 [1.1] | 64 / 22,949 [2.8] | 0.41 (0.25, 0.65) |
| Anaphylaxis | 0 / 24,019 [0] | 1 / 23,451 [0.043] | 0 (0, 38) |
| Nausea | 60 / 23,615 [2.5] | 112 / 22,613 [5] | 0.51 (0.37, 0.71) |
| Headache | 3 / 23,997 [0.13] | 1 / 23,465 [0.043] | 2.9 (0.24, 150) |
| Diarrhea | 28 / 23,773 [1.2] | 58 / 23,095 [2.5] | 0.47 (0.29, 0.75) |
| Lymphadenopathy | 14 / 23,898 [0.59] | 27 / 23,240 [1.2] | 0.67 (0.5, 0.88) |
| Fatigue | 91 / 23,224 [3.9] | 132 / 22,426 [5.9] | 2 (0.97, 4.2) |

**Table S12.** Incidence rates of adverse effects in the 21 days following the date of first vaccination for all individuals with at least one clinical note recorded during this time. For each adverse effect, incidence rates were calculated for the propensity matched vaccinated and unvaccinated individuals (n = 3,553 pairs) as the number of positive cases divided by the total number of at risk person days during this time period. Individuals were considered at risk for developing an adverse effect from their actual or assigned date of first vaccination until they experienced the event, died, or reached the end of the 21-day study period, or until four days prior to a positive SARS-CoV-2 test. For example, in the first row, we show that 100 cases of myalgia were recorded over a total of 72,586 person-days (contributed by 3,553 vaccinated individuals), corresponding to an incidence rate of 1.4 cases per 1000 person-days.

| **Adverse effect** | **Vaccinated**<br>**Incidence Rate**<br>Cases / Person-Days<br>*[Cases Per 1000*<br>*Person-Days]* | **Unvaccinated**<br>**Incidence Rate**<br>Cases / Person-Days<br>*[Cases Per 1000*<br>*Person-Days]* | **Incidence Rate**<br>**Ratio (95% CI)** |
| --- | --- | --- | --- |
| Myalgia | 100 / 72,586 [1.4] | 98 / 71,817 [1.4] | 1 (0.76, 1.3) |
| Fever | 55 / 73,085 [0.75] | 92 / 71,973 [1.3] | 0.59 (0.41, 0.83) |
| Chills | 32 / 73,364 [0.44] | 62 / 72,210 [0.86] | 0.51 (0.32, 0.79) |
| Facial paralysis | 4 / 73,660 [0.054] | 6 / 72,802 [0.082] | 0.66 (0.14, 2.8) |
| Erythema | 128 / 72,417 [1.8] | 155 / 71,319 [2.2] | 0.81 (0.64, 1) |
| Arthralgia | 141 / 72,063 [2] | 128 / 71,516 [1.8] | 1.1 (0.85, 1.4) |
| Vomiting | 112 / 72,569 [1.5] | 180 / 70,937 [2.5] | 0.61 (0.48, 0.77) |
| Anaphylaxis | 1 / 73,676 [0.014] | 1 / 72,882 [0.014] | 0.99 (0.013, 78) |
| Nausea | 226 / 71,499 [3.2] | 322 / 69,432 [4.6] | 0.68 (0.57, 0.81) |
| Headache | 7 / 73,616 [0.095] | 9 / 72,775 [0.12] | 0.77 (0.24, 2.3) |
| Diarrhea | 113 / 72,630 [1.6] | 178 / 71,176 [2.5] | 0.62 (0.49, 0.79) |
| Lymphadenopathy | 69 / 72,972 [0.95] | 89 / 71,888 [1.2] | 0.76 (0.55, 1.1) |
| Fatigue | 334 / 70,024 [4.8] | 437 / 68,245 [6.4] | 0.74 (0.64, 0.86) |

**Table S13.** Incidence rates of adverse effects in the 21 days following the date of second vaccination for all individuals with at least one clinical note recorded during this time. For each adverse effect, incidence rates were calculated for the propensity matched vaccinated and unvaccinated individuals (n = 2,360 pairs) as the number of positive cases divided by the total number of at risk person days during this time period. Individuals were considered at risk for developing an adverse effect from their actual or assigned date of first vaccination until they experienced the event, died, or reached the end of the 21-day study period, or until four days prior to a positive SARS-CoV-2 test. For example, in the first row, we show that 45 cases of myalgia were recorded over a total of 48,944 person-days (contributed by 2,360 vaccinated individuals), corresponding to an incidence rate of 0.92 cases per 1000 person-days.

| **Adverse effect** | **Vaccinated**<br>**Incidence Rate**<br>**Cases / Person-Days**<br>**[Cases Per 1000**<br>**Person-Days]** | **Unvaccinated**<br>**Incidence Rate**<br>**Cases / Person-Days**<br>**[Cases Per 1000**<br>**Person-Days]** | **Incidence Rate**<br>**Ratio (95% CI)** |
| --- | --- | --- | --- |
| Myalgia | 45 / 48,944 [0.92] | 36 / 48,039 [0.75] | 1.2 (0.77, 2) |
| Fever | 21 / 49,254 [0.43] | 35 / 48,050 [0.73] | 0.59 (0.32, 1) |
| Chills | 20 / 49,262 [0.41] | 34 / 48,018 [0.71] | 0.57 (0.31, 1) |
| Facial paralysis | 1 / 49,511 [0.02] | 1 / 48,448 [0.021] | 0.98 (0.012, 77) |
| Erythema | 39 / 49,052 [0.8] | 54 / 47,818 [1.1] | 0.7 (0.45, 1.1) |
| Arthralgia | 50 / 48,862 [1] | 49 / 47,819 [1] | 1 (0.66, 1.5) |
| Vomiting | 46 / 49,007 [0.94] | 96 / 47,243 [2] | 0.46 (0.32, 0.66) |
| Anaphylaxis | 0 / 49,527 [0] | 1 / 48,433 [0.021] | 0 (0, 38) |
| Nausea | 91 / 48,463 [1.9] | 167 / 46,423 [3.6] | 0.52 (0.4, 0.68) |
| Headache | 4 / 49,478 [0.081] | 3 / 48,423 [0.062] | 1.3 (0.22, 8.9) |
| Diarrhea | 50 / 48,901 [1] | 88 / 47,586 [1.8] | 0.55 (0.38, 0.79) |
| Lymphadenopathy | 19 / 49,291 [0.39] | 41 / 47,919 [0.86] | 0.45 (0.25, 0.79) |
| Fatigue | 122 / 47,823 [2.6] | 189 / 46,110 [4.1] | 0.62 (0.49, 0.79) |

**Table S14.** Comparison of adverse effect rates in the 7 days after each vaccine dose per solicited recording via V-Safe versus unsolicited documentation in EHR notes. V-Safe reporting data summarized through January 14, 2021 were extracted from a publicly available Advisory Committee on Immunization Practices (ACIP) powerpoint for mRNA-1273 (Moderna) and BNT162b2 (Pfizer/BioNTech). For the Moderna and Pfizer/BioNTech cohorts, the values shown are the percentages of patients who reported each symptom within 7 days of either the first dose (for those receiving the Moderna vaccine) or the first or second vaccine dose (for those receiving the Pfizer/BioNTech vaccine). Patient counts are not provided for these cohorts because only percentages were available in the source V-Safe dataset. At the time of this most recent V-Safe data summary, 2nd doses of the Moderna vaccine had not been administered and thus are not included. EHR documentation rates for each adverse effect were derived from Tables 3 and 4, which include the number of individuals who contributed at least one EHR note documenting the occurrence of the given symptom within 7 days of the first (Table 3) or second (Table 4) dose. To obtain the “Reporting to Documentation Ratio”, we divide the appropriate trial-derived reporting percentage by the corresponding EHR-derived documentation percentage.

|  | Moderna<br>(mRNA-1273, n = 1,083,174) | Pfizer/BioNTech<br>(BNT162b2, n = 997,042) |  | Mayo Clinic EHR<br>(mRNA-1273 + BNT162b2) |  | Reporting to Documentation Ratio<br>(Moderna / Mayo Clinic) | Reporting to Documentation Ratio<br>(Pfizer / Mayo Clinic) |  |
| --- | --- | --- | --- | --- | --- | --- | --- | --- |
| Adverse Effect | Dose 1 | Dose 1 | Dose 2 | Dose 1<br>(n = 31,609) | Dose 2<br>(n = 17,607) | Dose 1<br>[95% CI] | Dose 1<br>[95% CI] | Dose 2<br>[95% CI] |
| Fatigue | (29.7%) | (28.6%) | (50.0%) | 542<br>(1.7%) | 200<br>(1.1%) | 17 (16, 19) | 17 (15, 18) | 44 (38, 50) |
| Headache | (26.0%) | (25.6%) | (41.9%) | 12 (0.0%) | 7 (0.0%) | 680 (380, 1100) | 670 (370, 1100) | 1100 (480, 2000) |
| Myalgia | (19.6%) | (17.2%) | (41.6%) | 128<br>(0.4%) | 73 (0.4%) | 48 (41, 57) | 42 (36, 50) | 100 (79, 130) |
| Arthralgia | (8.6%) | (7.1%) | (21.2%) | 198<br>(0.6%) | 97 (0.6%) | 14 (12, 16) | 11 (9.8, 13) | 38 (31, 47) |
| Chills | (9.3%) | (7.0%) | (26.7%) | 50 (0.2%) | 38 (0.2%) | 59 (44, 77) | 44 (33, 58) | 120 (89, 170) |
| Nausea | (7.7%) | (7.0%) | (13.9%) | 321<br>(1.0%) | 143<br>(0.8%) | 7.6 (6.8, 8.4) | 6.9 (6.2, 7.7) | 17 (14, 20) |
| Fever | (9.1%) | (7.4%) | (25.2%) | 66 (0.2%) | 31 (0.2%) | 44 (34, 55) | 35 (28, 45) | 140 (99, 200) |

## References

1. CDC. COVID Data Tracker. https://covid.cdc.gov/covid-data-tracker/ (2020).

2. FDA authorizes Moderna COVID-19 vaccine. Med. Lett. Drugs Ther. 63, 9–10 (2021).

3. Office of the Commissioner. Pfizer-BioNTech COVID-19 Vaccine. https://www.fda.gov/emergency-preparedness-and-response/coronavirus-disease-2019-covid-19/pfizer-biontech-covid-19-vaccine (2021).

4. Baden, L. R. et al. Efficacy and Safety of the mRNA-1273 SARS-CoV-2 Vaccine. N. Engl. J. Med. 384, 403–416 (2021).

5. Polack, F. P. et al. Safety and Efficacy of the BNT162b2 mRNA Covid-19 Vaccine. N. Engl. J. Med. 383, 2603–2615 (2020).

6. [No title]. https://www.fda.gov/media/144413/download.

7. [No title]. https://www.fda.gov/media/144637/download.

8. CDC. CDC’s COVID-19 Vaccine Rollout Recommendations. https://www.cdc.gov/coronavirus/2019-ncov/vaccines/recommendations.html (2021).

9. Nguyen, L. H. et al. Risk of COVID-19 among front-line health-care workers and the general community: a prospective cohort study. Lancet Public Health 5, e475–e483 (2020).

10. Mutambudzi, M. et al. Occupation and risk of severe COVID-19: prospective cohort study of 120 075 UK Biobank participants. Occup. Environ. Med. (2020) doi:10.1136/oemed-2020-106731.

11. Panagiotou, O. A. et al. Risk Factors Associated With All-Cause 30-Day Mortality in Nursing Home Residents With COVID-19. JAMA Intern. Med. (2021) doi:10.1001/jamainternmed.2020.7968.

12. Adhikari, E. H. & Spong, C. Y. COVID-19 Vaccination in Pregnant and Lactating Women. JAMA (2021) doi:10.1001/jama.2021.1658.

13. Wu, K. Boston Doctor Reports Serious Allergic Reaction After Getting Moderna’s Covid Vaccine. New York Times (2020).

14. CDC COVID-19 Response Team, FDA. Allergic Reactions Including Anaphylaxis After Receipt of the First Dose of Moderna COVID-19 Vaccine — United States, December 21, 2020–January 10, 2021. (2021).

15. Covid, C. & Team, R. Allergic reactions including anaphylaxis after receipt of the first dose of Pfizer-BioNTech COVID-19 vaccine—United States, December 14--23, 2020. MMWR Surveill. Summ. 70, 46 (2021).

16. What to Know About Post-Vaccine Deaths and Allergies. The Washington Post (2021).

17. Castells, M. C. & Phillips, E. J. Maintaining Safety with SARS-CoV-2 Vaccines. N. Engl. J. Med. (2020) doi:10.1056/NEJMra2035343.

18. United States Department of Health and Human Services, Department of Health and Human Services (DHHS), Public Health Service (PHS), Food and Drug Administration (FDA) / Centers for Disease Control (CDC). Vaccine Adverse Event Reporting System (VAERS), CDC WONDER Online Database. (2021).

19. Pawlowski, C. et al. Pre-existing conditions are associated with COVID patients’ hospitalization, despite confirmed clearance of SARS-CoV-2 virus. medRxiv (2020).

20. Pawlowski, C. et al. Enoxaparin Is Associated With Lower Rates of Thrombosis, Kidney Injury, and Mortality Than Unfractionated Heparin in Hospitalized COVID Patients. (2020) doi:10.2139/ssrn.3707421.

21. Pawlowski, C. et al. Exploratory analysis of immunization records highlights decreased SARS-CoV-2 rates in individuals with recent non-COVID-19 vaccinations. medRxiv (2020).

22. Awasthi, S. et al. Plasma IL-6 Levels following Corticosteroid Therapy as an Indicator of ICU Length of Stay in Critically ill COVID-19 Patients. medRxiv (2020).

23. Agarwal, V. et al. Quantifying the prevalence of SARS-CoV-2 long-term shedding among non-hospitalized COVID-19 patients. medRxiv (2020) doi:10.1101/2020.06.02.20120774.

24. Pawlowski, C. et al. FDA-authorized COVID-19 vaccines are effective per real-world evidence synthesized across a multi-state health system. medRxiv (2021) doi:10.31219/osf.io/y6pdw.

25. Wagner, T. et al. Augmented curation of clinical notes from a massive EHR system reveals symptoms of impending COVID-19 diagnosis. Elife 9, (2020).

26. Devlin, J., Chang, M.-W., Lee, K. & Toutanova, K. BERT: Pre-training of Deep Bidirectional Transformers for Language Understanding. (2018).

27. The R Project for Statistical Computing. https://www.R-project.org/.

28. Sahai, H. & Khurshid, A. Statistics in Epidemiology: Methods, Techniques and Applications. (CRC Press, 1995).

29. Advisory Committee on Immunization Practices (ACIP), Tom Shimabukuro, MD, MPH, MBA. COVID-19 vaccine safety update. (2021).

30. Fernandez, M. A. L. migariane/DeltaMethodTutorial: Delta-method in Epidemiology. (2020) doi:10.5281/zenodo.3765172.

31. U.S. Food and Drug Administration. Moderna COVID-19 vaccine EUA fact sheet for health care providers. https://www.fda.gov/media/144637/download.

32. U.S. Food and Drug Administration. Pfizer-BioNTech COVID-19 vaccine EUA fact sheet for healthcare providers administering vaccine (vaccination providers). https://www.fda.gov/media/144413.

33. Jackson, L. A. et al. An mRNA vaccine against SARS-CoV-2—preliminary report. N. Engl. J. Med. (2020).

34. Mulligan, M. J. et al. Phase I/II study of COVID-19 RNA vaccine BNT162b1 in adults. Nature 586, 589–593 (2020).

